# Proteome-wide antibody responses to PfSPZ Vaccine include cross-reactive antibodies that correlate with protection from malaria

**DOI:** 10.64898/2026.11.16.26367676

**Authors:** Andrea A. Berry, Olukemi O. Ifeonu, DeAnna J. Friedman-Klabanoff, Amit Oberai, Adam D. Shandling, Joshua M. Edgar, Jozelyn V. Pablo, Andy A. Teng, Mark A. Travassos, Philip L. Felgner, Fadil A. Bidmos, Richard S. Pinapati, John C. Tan, Christopher V. Plowe, Judith E. Epstein, Thomas L. Richie, Robert A. Seder, Stephen L. Hoffman, Shannon Takala-Harrison, Kirsten E. Lyke, Azza H. Idris, Joseph J. Campo

## Abstract

The PfSPZ Vaccine induces both humoral and cellular immunity. While T cells are likely critical for protection, previous analyses of antibody correlates have focused on select antigens. Here, we apply a proteome-wide approach to examine antibody responses across the entire *Plasmodium falciparum* (Pf) proteome. Sera from malaria-naïve volunteers immunized with high doses of PfSPZ Vaccine were probed using whole proteome Pf protein microarrays and high-density Pf peptide microarrays. Both platforms identified antibody responses that significantly correlated with protection, including responses to the immunodominant Pf circumsporozoite protein (CSP). Multiple reactive epitopes within non-CSP proteins shared homology with the central repeat and junctional regions of CSP, suggesting broad intra-proteome cross-reactivity. Greater breadth of seropositive responses was associated with protection. Cross-reactivity was confirmed by off-target binding of well-characterized memory B cell- and plasmablast-derived monoclonal antibodies (mAbs) targeting CSP. What began as a search for antibody correlates of protection and potential vaccine candidate antigens led to the surprising discovery of probable promiscuous CSP antibody responses that bind non-CSP proteins. The association between protection and greater levels of intra- and extra-*Plasmodium* cross-reactivity raises new questions about parasite biology and microbial evolution.

## INTRODUCTION

Despite decades of eradication efforts, malaria remains a major cause of global morbidity and mortality. The largest malaria burden is caused by *Plasmodium falciparum* (Pf) and predominantly affects children in sub-Saharan Africa (1). Vaccination is seen as one of the most important tools for controlling malaria, but vaccine development has historically been hampered by the complex multi-stage life-cycle and genomic diversity of the parasite (2). After decades of clinical development, the RTS,S/AS01E and R21/Matrix-M malaria vaccines have been recommended by the World Health Organization (1). Both are subunit vaccines based on the Pf circumsporozoite protein (CSP), the immunodominant surface protein expressed on the pre-erythrocytic stage sporozoites. Immune correlates and mechanisms of protection for these vaccines are likely through induction of anti-CSP antibodies (3).

Another pre-erythrocytic vaccine approach is immunization with whole-organism radiation-attenuated Pf sporozoites, which has achieved very high levels of protection in clinical studies (4–7). The most advanced PfSPZ vaccine is Sanaria® PfSPZ Vaccine, comprised of radiation attenuated, early arresting, non-replicating, aseptic, purified, vialed, cryopreserved PfSPZ (8). PfSPZ Vaccine has been safe and efficacious at preventing Pf infection in malaria naïve recipients and in recipients living in malaria endemic areas. Protection elicited by PfSPZ Vaccine is thought to be mediated largely by tissue resident cells, particularly CD8+ T cells. However, protection against Pf infection in the field for 12 to 77 weeks after the last immunization has been correlated at the population level with anti-CSP antibodies (9–11). Unlike the RTS,S and R21 vaccines, which include only a single Pf protein, metabolically active PfSPZ may present any of hundreds to a thousand parasite antigens (12). After Pf sporozoites invade hepatocytes, parasite development continues for 5-6 days, during which infected hepatocytes can be cleared by cellular immune responses toward MHC-peptide complexes (13). Beyond CSP, antibodies to a small number of liver and blood stage antigens have been identified, but the antigens studied were chosen a priori rather than through an unbiased, proteome-wide screen (14). Identification of Pf proteins associated with antibody responses that correlate with protection may accelerate discovery of predictive markers of vaccine efficacy, or may themselves be candidates for development as next generation pre-erythrocytic vaccines.

Recent proteomic antibody profiling studies in the infants and children who participated in the RTS,S/AS01E phase 3 trial showed that a CSP-only subunit vaccine can elicit broad off-target antibody responses to non-CSP Pf proteins (15). It remains unclear if these antibodies bind to cross-reactive epitopes, if they are the result of boosted pre-existing antibodies, or if they are a primary antibody response associated with parasite killing during ongoing exposure. Homology analysis identified NANP-like motifs on non-CSP proteins, suggesting cross-reactive binding by CSP NANP-elicited antibodies; however, many post-RTS,S antibody responses target proteins that lack NANP-like epitopes. In a separate cohort of malaria-naïve adults vaccinated with RTS,S and challenged by CHMI, in whom boosting and ongoing exposure can be excluded, off-target IgG and IgA responses to 95% of non-CSP proteins studied were higher in protected than in unprotected vaccinees, and 22 differentially reactive regions shared sequence similarity with CSP epitopes (16). In both studies, higher “off-target” responses were associated with protection. Confirming whether these responses represent cross-reactive CSP antibodies or protein-specific responses informs whether the association with protection reflects a surrogate of vaccine immunogenicity or suggests protective mechanisms involving non-CSP antigens.

To comprehensively analyze the PfSPZ antibody response following vaccination with PfSPZ Vaccine and identify correlates of protection, serum samples from participants who received the PfSPZ Vaccine, and were either protected or not protected following controlled human malaria infection (CHMI), were analyzed using two independent proteomic antibody-profiling platforms: protein microarrays and high-density peptide microarrays. A focus on malaria naïve adults who received high doses of PfSPZ Vaccine provides a high-resolution view of the proteome-wide antibody response to sporozoites and early liver stage development. The goal was to define the breadth and nature of the Pf proteins recognized after PfSPZ vaccination at both the protein and epitope levels, and to clarify whether previously observed anti-CSP binding to non-CSP proteins represents CSP-driven cross-reactivity.

## RESULTS

### PfSPZ Vaccine volunteers and proteomic microarrays selected for analysis

To identify correlates of protection, serum samples were selected from three clinical trials of PfSPZ Vaccine in malaria-naïve adults: VRC-312 (17), VRC-314 (18) and Warfighter 1 (WRAIR-2080) (14). To maximize the power to detect differences between the protected and the not protected groups, groups from previous studies were combined for this analysis. The included groups received ≤ 4 doses of ≥ 1.35 x 10^5^ sporozoites per dose by DVI or IM, underwent CHMI 3 weeks after the final immunization, and included both protected and not-protected participants. For each participant, sera collected at baseline (pre-vaccination) and prior to CHMI (3 weeks after the final vaccination, i.e., post-vaccination) were probed on both full-proteome protein microarrays and high-density peptide microarrays. The group receiving 4 doses of 2.2 x 10^5^ PfSPZ IM was assessed only on protein microarrays. Study sample details are shown in **Table 1**. The protein microarrays contained 8,871 full-length or fragmented Pf proteins expressed using a coupled cell-free *E. coli* in vitro transcription and translation (IVTT) system, representing 5,233 unique Pf 3D7 strain genes (3D7 is a clone of PfNF54, the parasite strain used for development of PfSPZ Vaccine). The high-density peptide microarrays consisted of 16-mer peptides synthesized by light-directed solid-phase peptide synthesis, each peptide overlapping by 15 amino acids with coverage of 5,362 genes (6,060 inclusive of isoforms) from the NF54 strain proteome (total of 320,909 peptides).

**Table 1.**
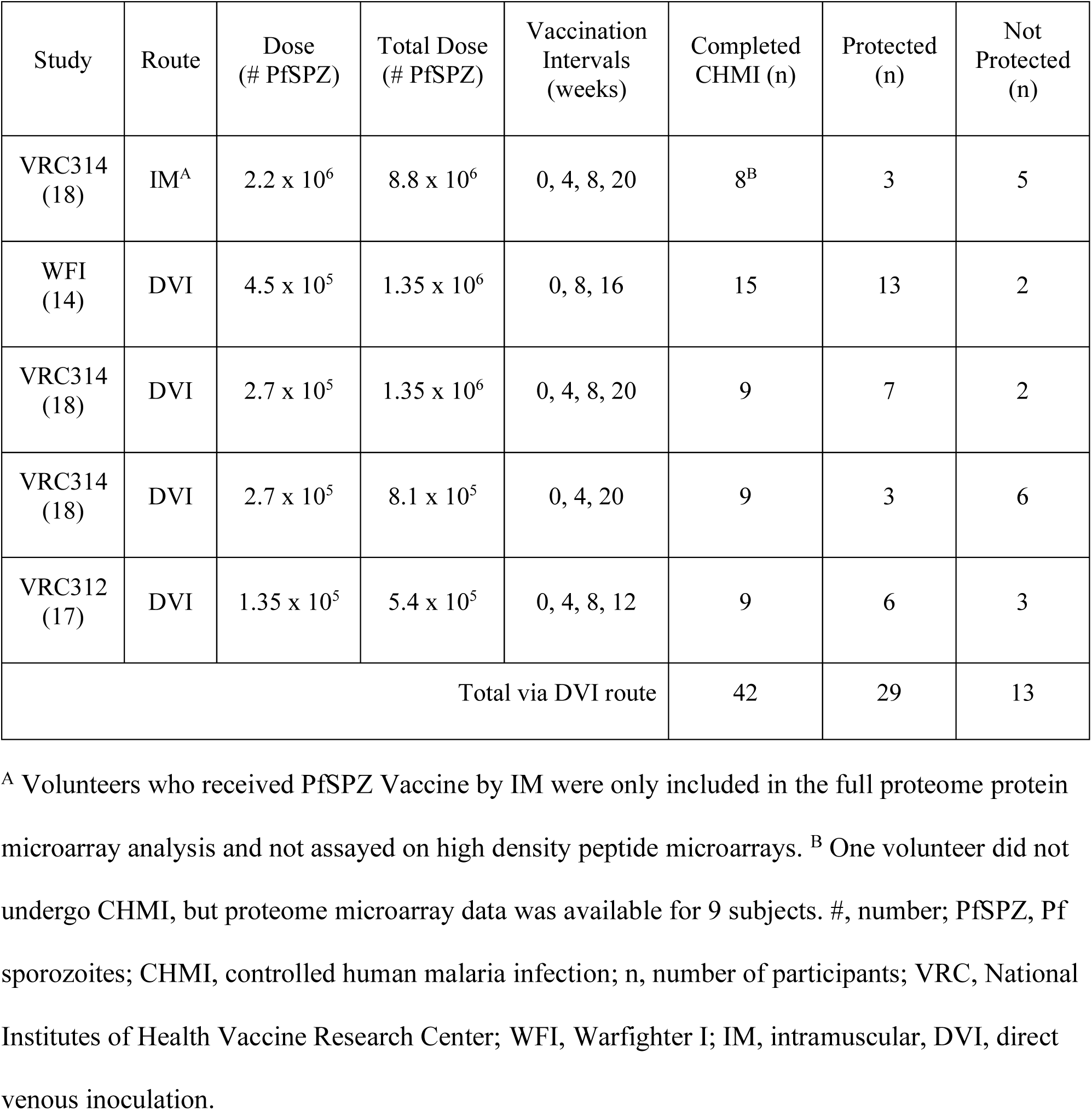
Origin of samples from completed PfSPZ Vaccine Clinical Trials in the current study.

| Study | Route | Dose (# PfSPZ) | Total Dose (# PfSPZ) | Vaccination Intervals (weeks) | Completed CHMI (n) | Protected (n) | Not Protected (n) |
|---|---|---|---|---|---|---|---|
| VRC314 (18) | IM<sup>A</sup> | 2.2 x 10<sup>6</sup> | 8.8 x 10<sup>6</sup> | 0, 4, 8, 20 | 8<sup>B</sup> | 3 | 5 |
| WFI (14) | DVI | 4.5 x 10<sup>5</sup> | 1.35 x 10<sup>6</sup> | 0, 8, 16 | 15 | 13 | 2 |
| VRC314 (18) | DVI | 2.7 x 10<sup>5</sup> | 1.35 x 10<sup>6</sup> | 0, 4, 8, 20 | 9 | 7 | 2 |
| VRC314 (18) | DVI | 2.7 x 10<sup>5</sup> | 8.1 x 10<sup>5</sup> | 0, 4, 20 | 9 | 3 | 6 |
| VRC312 (17) | DVI | 1.35 x 10<sup>5</sup> | 5.4 x 10<sup>5</sup> | 0, 4, 8, 12 | 9 | 6 | 3 |
| Total via DVI route |  |  |  |  | 42 | 29 | 13 |

<sup>A</sup> Volunteers who received PfSPZ Vaccine by IM were only included in the full proteome protein microarray analysis and not assayed on high density peptide microarrays. <sup>B</sup> One volunteer did not undergo CHMI, but proteome microarray data was available for 9 subjects. #, number; PfSPZ, Pf sporozoites; CHMI, controlled human malaria infection; n, number of participants; VRC, National Institutes of Health Vaccine Research Center; WFI, Warfighter I; IM, intramuscular, DVI, direct venous inoculation.

### The proteome-wide antibody response to PfSPZ Vaccine

Among the 42 volunteers who received PfSPZ Vaccine by DVI and the 9 volunteers who received it by IM injection, pre- and post-vaccination antibody profiles demonstrated substantial heterogeneity both between individuals and across proteins (**Fig. 1, S1**). Antibody signals measured on the protein microarray were compared with the negative signal distribution to determine serorecognition; for the whole-proteome analyses, most proteins are assumed to be non-reactive to immunoglobulins, and therefore the median signal of all protein spots serves as background. A total of 241 of the 8,871 Pf proteins on the protein microarray were serorecognized, defined as normalized IgG values ≥ 1.0 (twice the background) in at least 50% of post-vaccination samples within any vaccine regimen. These full length or fragmented Pf proteins represented 220 unique Pf genes (**Fig. 1A, Table S1**). By comparison, only 41 protein microarray proteins were recognized at the pre-vaccination timepoint (**Fig. S2**). For the peptide microarray analysis, data were first collapsed from the peptide level to the protein level by using the peak antibody signal for each protein-sample pair. Serorecognized proteins were those where post-vaccination antibody response > mean + 2.5 standard deviations of all pre-vaccination antibody responses for at least 50% of post-vaccination samples. A total of 69 of 6,060 proteins on the peptide microarray, representing 67 unique Pf proteins, were serorecognized (**Table S2**). The distribution of IgG responses to the features on the protein and peptide microarrays was similar across all vaccine regimens (**Fig. 1B-C**). When performing unsupervised dimension reduction and projection using tSNE on all protein microarray proteins, samples from subjects did not cluster by vaccine regimen, but tended to project closest or in close proximity to samples from the same subject, representing a “fingerprint” antibody profile that is unaffected by immunization with PfSPZ Vaccine (**Fig. 1D**). In contrast, when reducing the data to only the 241 serorecognized proteins on the protein microarrays, a clear pattern emerged with post-vaccination samples clustered directionally away from pre-vaccination samples, representing a consistent effect of PfSPZ Vaccine on a subset of Pf proteins and not the whole proteome, i.e., immunogenicity (**Fig. 1E**). This pattern of both a whole proteome fingerprint and immunogenicity against a defined set of Pf proteins was consistent when varying the perplexity parameter of tSNE models (**Fig. S3A-F**), and when applying UMAP to project greater global structure of the data vs. the local structure-preserving nature of tSNE (**Fig. S3G-H**). Unsupervised dimension reduction and projection using tSNE on all peptide microarray peptides, as well as serorecognized peptides, also produced a “fingerprint” antibody profile with a few outliers present (**Fig. 1F-G**).

**Fig. 1.**
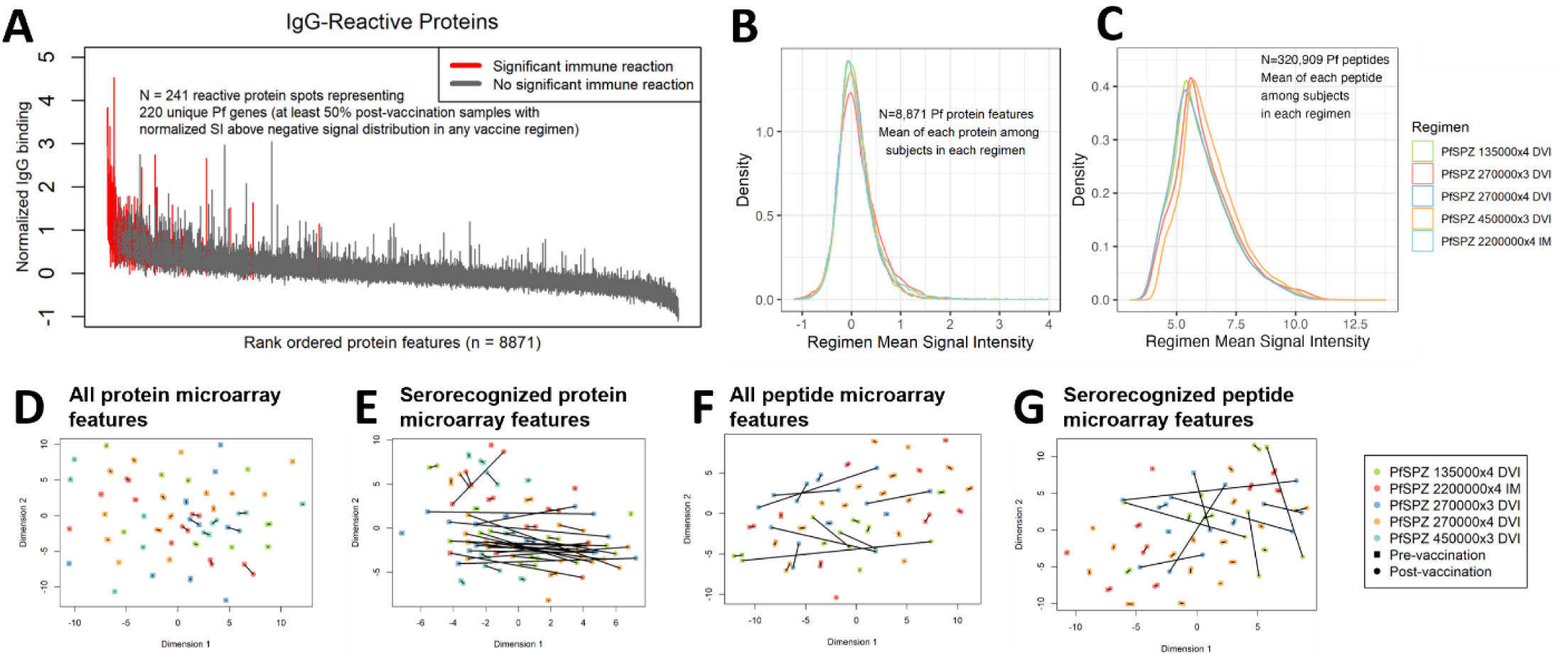
Proteome-wide IgG antibody reactivity to PfSPZ Vaccine. **(A)** The error bar plot shows the interquartile range of IgG seroreactivity to each protein feature (full-length or fragmented proteins) on the *P. falciparum* protein microarray. Protein features are ordered by median of all post-vaccination signals (global median), and bars are colored red if at least 50% of the study subjects in one or more vaccine regimens had positive responses to the protein (normalized signal intensity [SI] ≥ 1.0). **(B-C)** The density plots of the protein microarray and peptide microarray signals, respectively, are shown as density curves of the means for each protein or peptide for each PfSPZ Vaccine regimen. **(D-G)** t-Stochastic Neighbor Embedding (tSNE) projections of the data from all protein microarray proteins, only serorecognized antigens (red bars in **A**), all peptide microarray peptides, and peptides of only the serorecognized proteins, respectively. Points for each sample are colored by PfSPZ Vaccine regimen and paired pre-vaccination (square points) and post-immunization (round points) data are connected by black lines. The perplexity parameter used in protein microarray models was 30 and in peptide microarray models was 27.

To define the set of immunogenic Pf proteins associated with PfSPZ vaccination, longitudinal antibody levels measured on protein microarrays were modeled using linear mixed effects regression (LMER), controlling for vaccine regimen. Among the serorecognized protein microarray proteins, most showed an increasing trend following immunization by DVI, and antibody responses to 33 Pf proteins representing 31 unique genes were significantly increased after correction for the false discovery rate (FDR) (**Fig. 2A**). These included CSP and merozoite protein 5 (MSP5), which has been previously identified as a potential vaccine candidate antigen (19–23), but also novel proteins such as claudin-like apicomplexan microneme protein (CLAMP), double C2-like domain-containing protein (DOC2), leucine rich repeat protein 9 (LRR9), glycogen synthase kinase 3 (GSK3), surface-associated interspersed protein 8.1 (SURF8.1), a ring zinc finger protein (RZnFing) and other uncharacterized proteins, a subset of which were also identified in participants who had received RTS,S vaccine (24, 15). Interestingly, IM administration of PfSPZ Vaccine elicited a similar profile of antibodies with a more limited but overlapping set of antigens, except for thrombospondin-related anonymous protein (TRAP, also known as surface sporozoite protein 2 “SSP2”) and apical membrane antigen 1 (AMA1), which were uniquely recognized after IM administration (**Fig. 2B**). Similar results were obtained using paired T tests and Wilcoxon’s sign rank tests (**Table S3**). To validate the antibody immunogenicity signature, protein microarray responses to CSP and MSP5 were compared against independently performed ELISA, which showed a correlated linear relationship (**Fig. S4**).

**Fig. 2.**
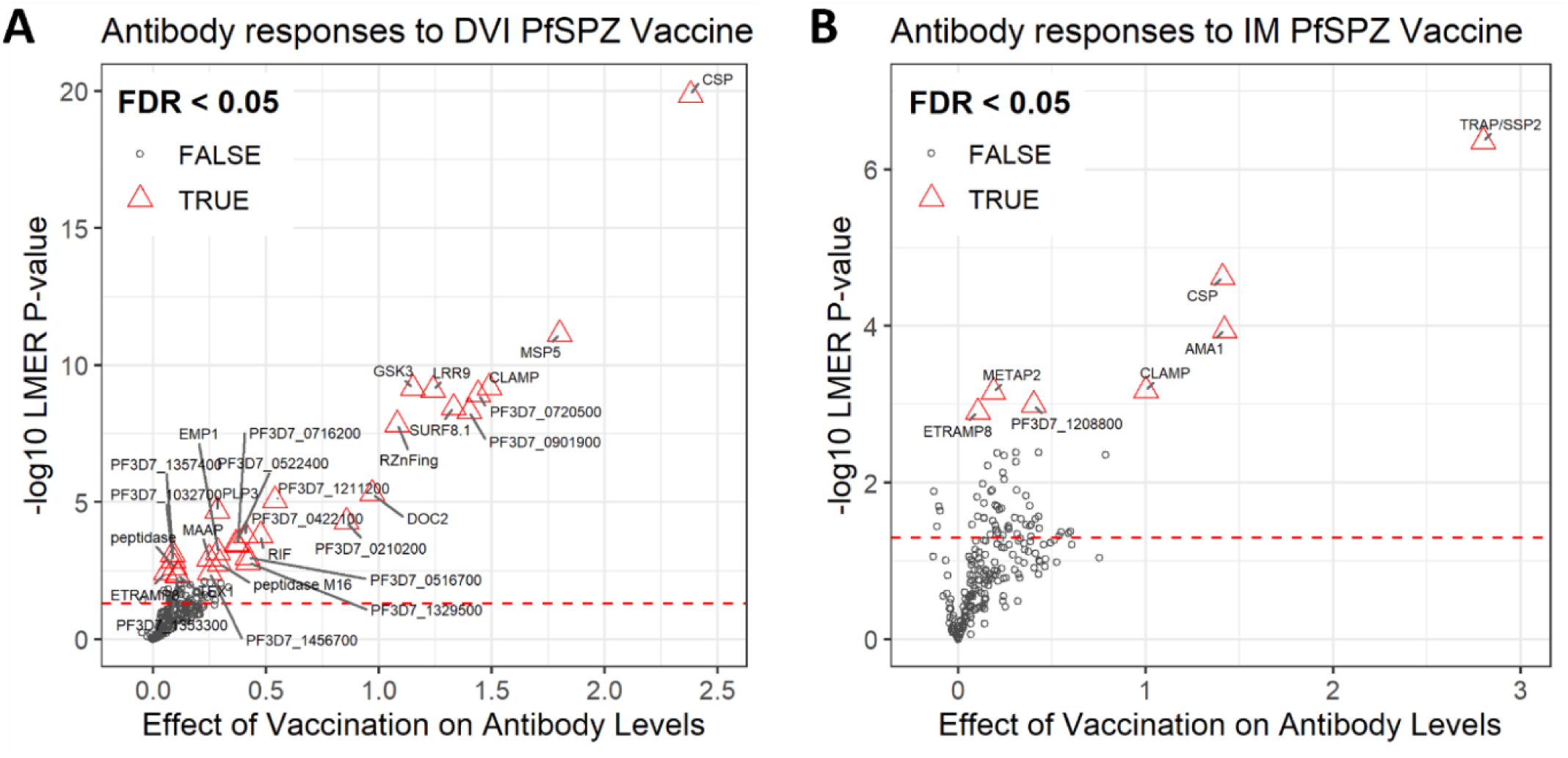
Immunogenicity of *P. falciparum* proteins after PfSPZ Vaccine delivered by direct venous inoculation (DVI) or intramuscular injection (IM). The volcano plots show the effect of vaccination on antibody levels as linear mixed effects regression (LMER) fixed effects coefficients (x-axis) and the inverse base-10 logarithm of LMER P-values calculated by full and null model likelihood ratio tests (y-axis) for each of the 241 reactive protein spots on protein microarrays (points) identified in Fig. 1A. Antigens are plotted with unadjusted P-values, and antibody responses that remain significant after correction for the false discovery rate (FDR) are shown as red labeled triangles. The red dashed lines represent unadjusted P-value of 0.05. **(A)** PfSPZ Vaccine regimens administered by direct venous injection (DVI, N=42 subjects, N=84 samples); **(B)** administered intramuscularly (IM, N=8 subjects, N=16 samples).

### Antibodies targeting CSP and multiple other Pf proteins are associated with protection from PfSPZ Challenge

To test which immunogenic proteins were associated with protection, permutation tests were performed between protected and non-protected individuals. For the protein microarrays, the change in antibody responses from pre-vaccination to post-vaccination (Delta) for each antibody response was compared. Antibodies significantly associated with protection after correction for the FDR included MSP5, RZnFing, CSP, SURF8.1, GSK3, perforin-like protein 3 (PLP3) and PF3D7_1211200 conserved protein of unknown function, with borderline significance for LRR9 and CLAMP (**Fig. 3A-B, Table S4**). There were no antigens significantly associated with protection in the IM vaccination regimen, although this result was expected due to the small number of individuals (**Fig. S5**). For the peptide microarrays, post-vaccination antibody levels were compared using permutation testing. Of the 67 serorecognized proteins on peptide microarrays, 35 proteins had antibody responses that were associated with protection (**Fig. 3C-D, Table S2**). For the 35 proteins associated with protection on the peptide microarray, putative epitopes (amino acid peaks) were identified as series of at least four consecutive amino acids for which the mean antibody signal in the protected group exceeded the serorecognition cutoff. An average of 1.8 peaks (median 1 peak) were identified per protein, with 15 proteins having multiple peaks (range 2-10 peaks). Signal intensity peaks for CSP, CLAMP, MSP5, and GSK3 are shown in **Fig. 4**.

**Fig. 3.**
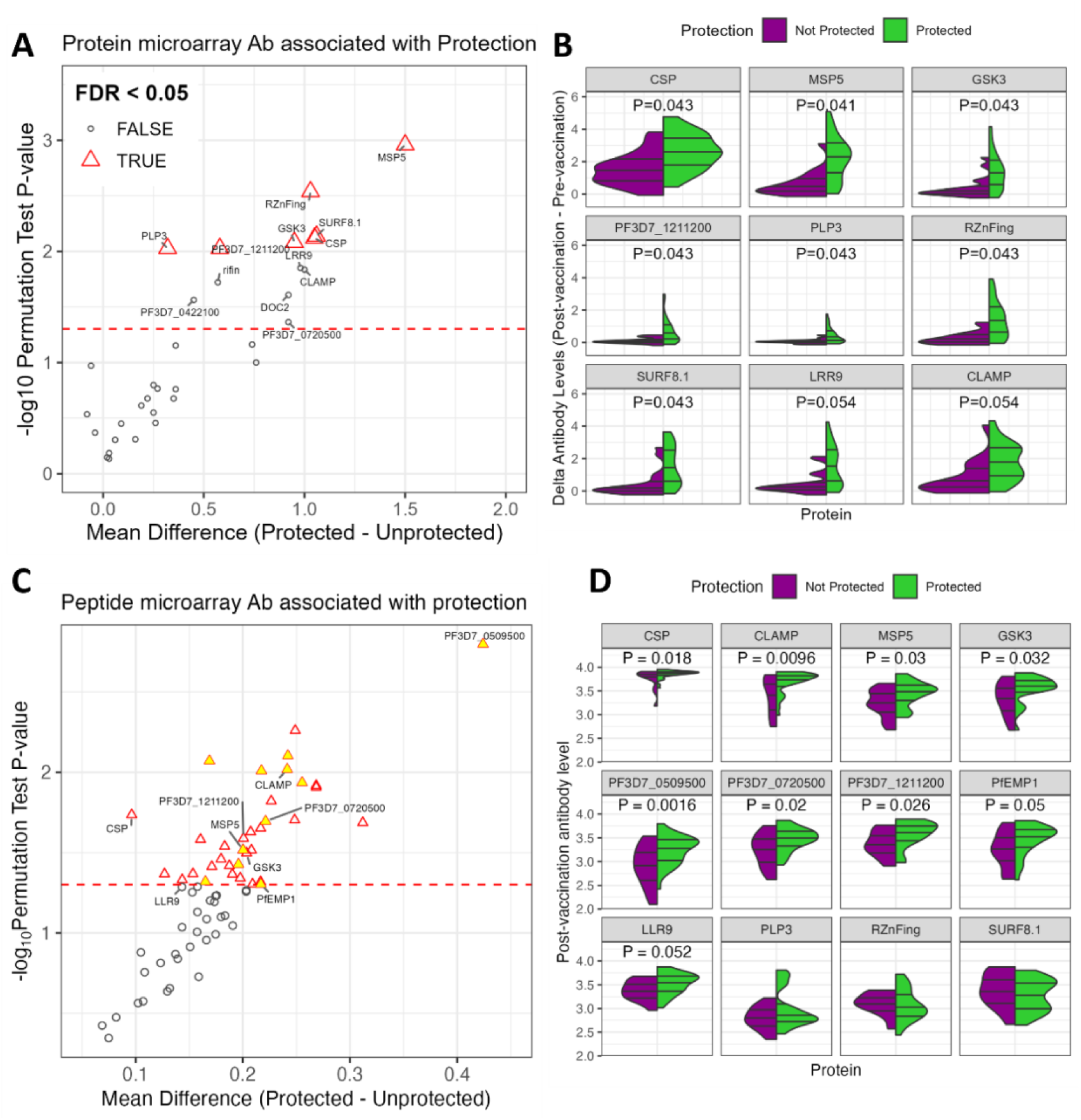
Antibodies associated with protection from PfSPZ Challenge. **(A)** The volcano plot shows the mean difference of the antibody Deltas (post-vaccination minus pre-vaccination signal intensity) between protected and unprotected subjects probed on protein microarrays (x-axis) by the inverse log10 P-value of permutation tests (y-axis) for each immunogenic antigen (points) identified in Fig. 2A. The dashed red line represents an unadjusted P-value of 0.05, and points shown as red labeled triangles represent antibody responses that remain significant after correction for the false discovery rate (FDR). **(B)** The split violin plots show the distribution of Delta antibody levels against 9 of the most significant antigens in (A) between protected (green) and not protected (purple) subjects. Lines within the violins represent the median and interquartile ranges. FDR-adjusted P-values from permutation tests are shown at the top of each panel. **(C)** The volcano plot shows the mean difference of the post-vaccination antibody levels between protected and unprotected subjects probed on peptide microarrays (x-axis) by the inverse log10 P-value of permutation tests (y-axis) for each of the 67 serorecognized proteins on peptide microarrays (points). The dashed red line represents an unadjusted P-value of 0.05, and points shown as red labeled triangles represent significant antibody responses (unadjusted P-values). Of the proteins with significant antibody responses, those containing reactive epitopes that aligned with CSP are colored yellow. **(D)** The split violin plots represent 12 of the most significant peptide microarray responses and/or proteins that were identified on protein microarrays, including some that were not significantly associated with protection (PfEMP1, LRR9) and some that did not have a post-vaccination response above the threshold value defined by the baseline samples (PLP3, RZnFing, SURF8.1).

**Fig. 4.**
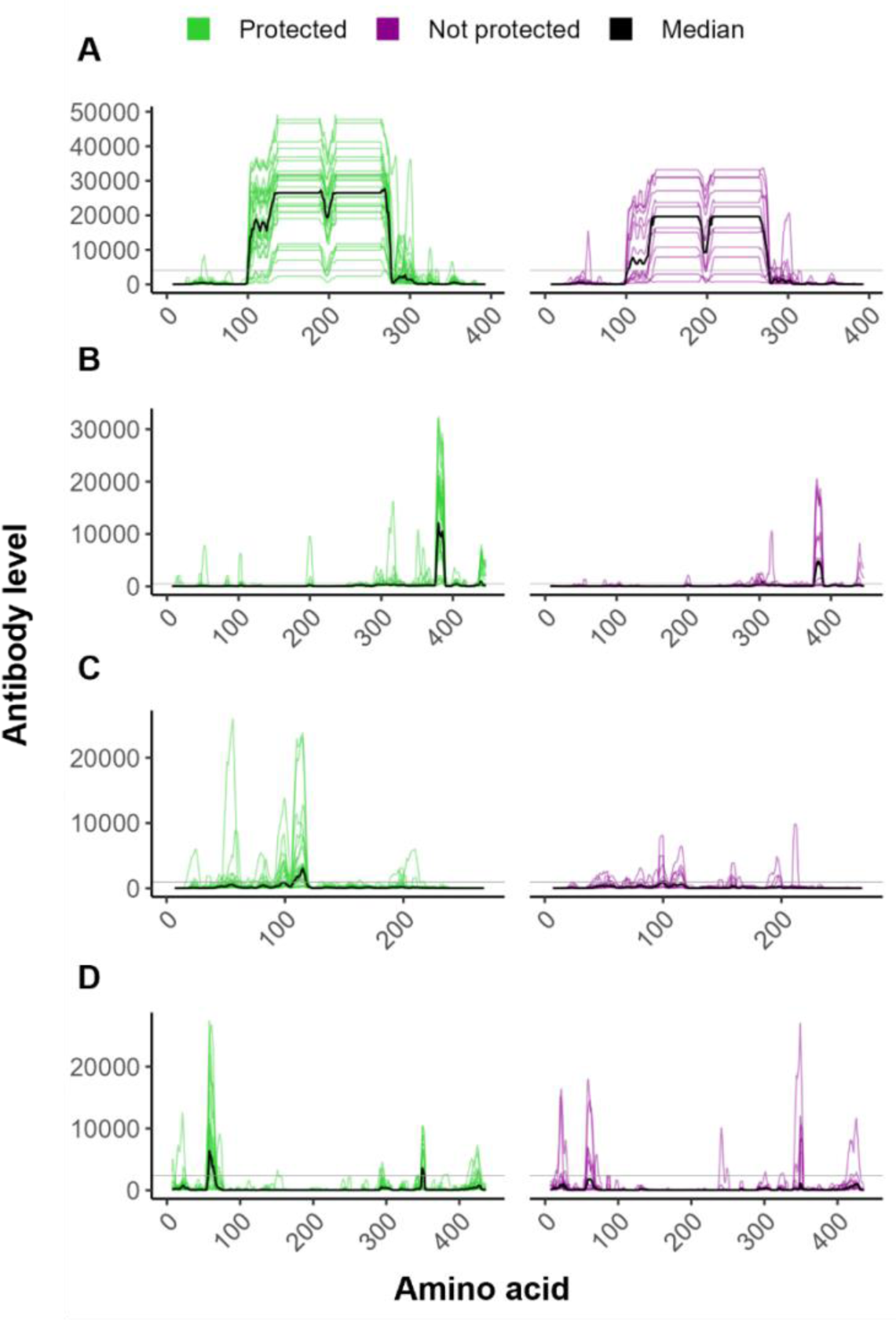
Peptide microarray antibody binding profiles against CSP, CLAMP, MSP5 and GSK3. Post-vaccination peptide microarray array seroreactivity to (**A**) PF3D7_0304600 (CSP), (**B**) PF3D7_1030200 (CLAMP), (**C**) PF3D7_0206900 (MSP5), and (**D**) PF3D7_0312400 (GSK3) are shown for each subject (thin lines) and as group medians (black lines) with signal intensity (y-axis) at each amino acid position (x-axis). Samples are shown as green lines for participants protected from PfSPZ Challenge and purple lines for not-protected participants.

We compared fold-change in antibody response (post-pre antibody Delta) for the 33 immunogenic proteins on the protein microarray. When subjects were grouped by antibody Deltas using hierarchical clustering, the two largest clusters separated into high responders and low-medium responders (**Fig. S6, Table S5**), which were associated with protection by logistic regression (*P*=0.029, *P_adj_*=0.068 adjusting for vaccine regimen). However, a group of medium-high responders were separated into several smaller trees that were not captured using unsupervised clustering. To show the association between having antibody responses to multiple antigens and protection, antibody “immunogenicity” counts were calculated as the sum of responses at least 2-fold over pre-vaccination levels per subject among the immunogenic proteins on protein microarrays (**Fig. 5A**). Higher breadth scores were significantly associated with protection (Wilcoxon rank sum test *P*=0.0067, **Fig. 5B**). Notably, antibody immunogenicity counts tended to increase in parallel with CSP antibody responses and were positively correlated with CSP antibody Deltas (Pearson’s correlation coefficient: 0.83; **Fig. 5C**). When assessing protective antibody Deltas by logistic regression controlling for the CSP response, all antigens independently were no longer significantly associated with protection. However, CSP levels were not always in concordance with antibody immunogenicity counts, with high CSP levels paired with low counts and vice-versa, suggesting that some protection-associated antibody responses are independent of CSP. Correlation of all immunogenic protein microarray antibody levels with CSP showed that a subset of proteins was positively correlated with CSP responses at varying levels, and another set of proteins was not, suggesting that the subset of correlated proteins may be related independently by collinearity or possibly be dependent on cross-reactive antibody binding epitopes (**Fig. 5D**).

**Fig. 5.**
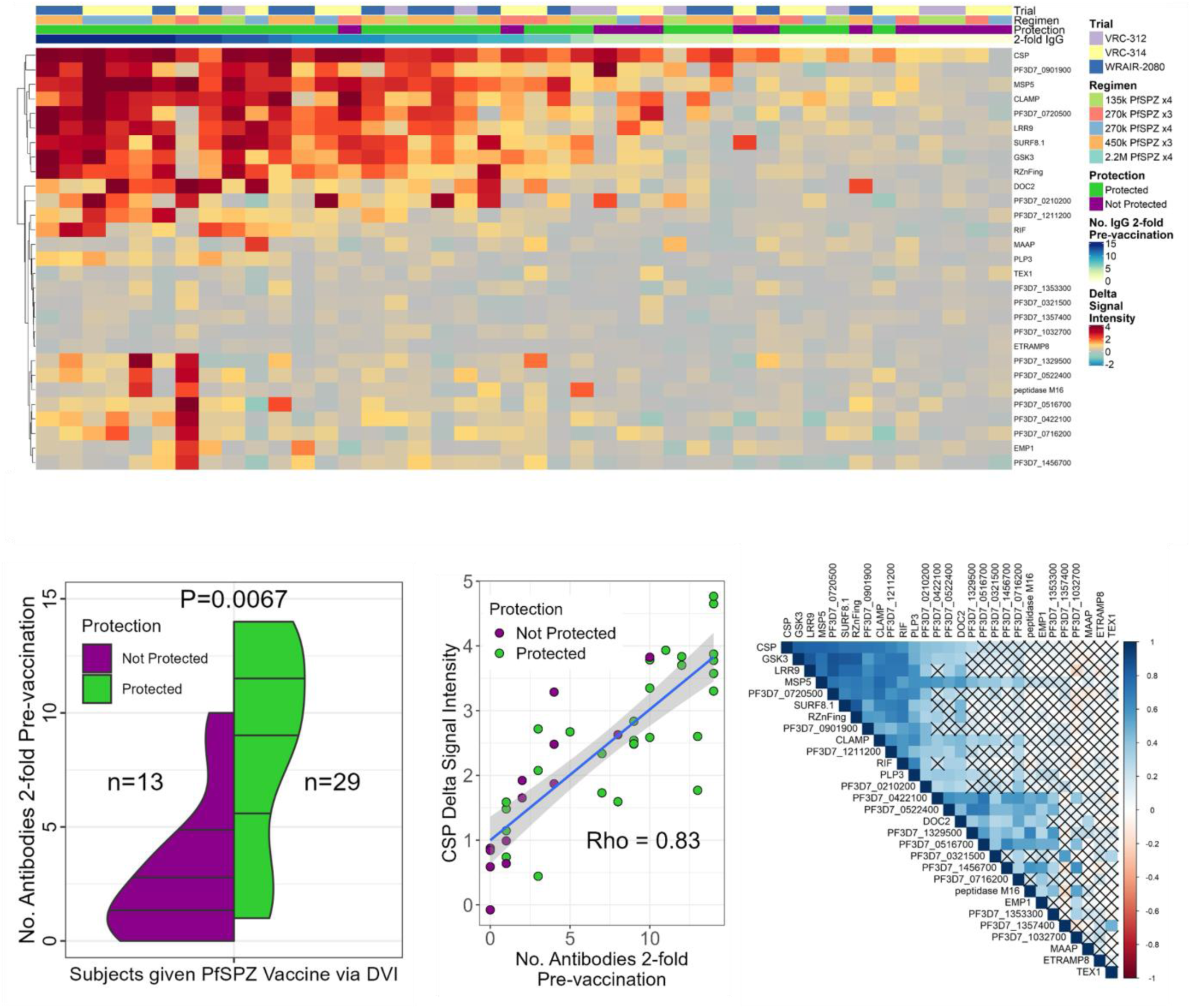
The number of antibody responses to PfSPZ Vaccine is associated with protection from PfSPZ Challenge and correlates with CSP antibodies. **(A)** The heat map displays the antibody Deltas of protein microarray signal intensity (post-vaccination minus pre-vaccination) on a grey to red scale for the 33 proteins identified as immunogenic on protein microarrays (Fig. 2A). Proteins (rows) are ordered by hierarchical clustering. Subjects (columns) are ordered by decreasing “antibody immunogenicity counts”, defined as the sum of responses that were 2-fold of IgG signal intensity in the post-vaccination samples over pre-vaccination samples (shown in white to blue scale in column headers). Protein labels are shown at the right of the heat map rows. Column labels indicate the trial, PfSPZ Vaccine regimen and protection status of each subject. **(B)** The split violin plot shows the distribution of the antibody immunogenicity counts (y-axis) among subjects who were not protected and protected from PfSPZ Challenge (purple and green, respectively) with associated Wilcoxon’s rank-sum test P-value. **(C)** The scatter plot shows protected (green) and not protected (purple) individuals’ Delta responses to the CSP (y-axis) vs. antibody immunogenicity counts (x-axis). Pearson’s correlation coefficient (Rho) is shown as a best fit line in blue and confidence band in grey. (D) The correlogram shows the Pearson’s correlation coefficients between antigen Deltas on a red to white to blue color scale for increasing correlation. Tiles with “X” over them indicate non-significant tests, i.e., no correlation.

### Epitope alignment identifies epitopes in multiple Pf proteins that may cross-react with CSP-targeting antibodies

BLAST-p was used for pairwise alignment of the putative epitopes (examples in **Fig. 4**) identified from peptide microarrays with the CSP (NF54) amino acid sequence. Putative epitopes from eleven proteins had sequence homology with CSP (**Table 2**). The putative epitopes aligned to CSP at the junctional region (n=4), the NVDP minor repeat motif of the central repeat region (n=1), the NANP major repeat motif of the central repeat region (n=2), the end of the central repeat region (n=3), and/or the beginning of the C-terminal region (N-terminal to the Th2R epitope, n=1) (**Fig. S7**). The sequence homology between the putative epitopes and CSP raises the possibility that the antibodies were elicited by CSP epitopes and then were cross-reactive with non-CSP peptides, but another possibility, given that PfSPZ Vaccine presents multiple proteins, is that the antibody reactivity to non-CSP proteins was elicited by distinct proteins.

**Table 2.** Putative cross-reactive *Plasmodium falciparum* circumsporozoite epitopes and locations of best alignment.

| Protein ID | Protein Name | Best alignment* | CSP location | PfSPZ Vaccinees | mAb4 | mAb10 |
|---|---|---|---|---|---|---|
| PF3D7_0206900 | merozoite surface protein-5 | NSNPNLD<br>NANPNVD | Junctional/NVDP | 1 |  | 1 |
| PF3D7_0513200 | CPSF domain-containing protein, putative | NGNPNI<br>NANPNV | Junctional/NVDP | 1 | 1 | 1 |
| PF3D7_1030200 | claudin-like apicomplexan microneme protein, putative | HMGPNLNPNFFP<br>NVDPNANPNVDP | Junctional | 1 | 1 | 1 |
| PF3D7_0723600 | proteasome assembly chaperone 4, putative | SNPNVD<br>ANPNVD | Junctional/NVDP | 1 |  |  |
| PF3D7_1355700 | NLI interacting factor-like phosphatase, putative | DTHVNTNPNF<br>NANPNANPNV | NVDP | 1 |  |  |
| PF3D7_0300100 | erythrocyte membrane protein 1, PfEMP-1 | GPNESPNSKP<br>NPNANPNANP | NANP | 1 |  |  |
| PF3D7_0509500 | ERCC4 domain-containing protein, putative | PNANPH<br>PNANPN | NANP | 1 |  |  |
| PF3D7_0720500 | conserved Pf protein, unknown function | NINPNQ<br>NANPNK | CRR end | 1 |  |  |
| PF3D7_1202300 | dynein heavy chain, putative | NANPNK<br>NANPNK | CRR end | 1 |  |  |
| PF3D7_1417600 | ookinete surface and oocyst capsule protein | PNASPNK<br>PNANPNK | CRR end | 1 |  |  |
| PF3D7_0727100 | conserved Pf protein, unknown function | PNDPNR<br>PNEPNW | C terminal | 1 |  |  |
| PF3D7_0809200 | asparagine-rich antigen Pfa55-14 | DIQVNPNKN<br>NPNANPNKN | NANP |  | 1 |  |
| PF3D7_0823300 | histone acetyltransferase GCN5 | MNPNMN<br>ANPNAN | NANP |  | 1 |  |
| PF3D7_1103800 | CCR4-NOT transcription complex NOT1-G, putative | PFNPNLKIDE<br>PNDPNRNVDE | CRR end |  | 1 |  |
| PF3D7_1335900 | thrombospondin-related anonymous protein | NPEEGKGENPNG<br>NPNANPNANPNA | NANP |  | 1 |  |
| PF3D7_0812100 | proteasome activator complex subunit 4, putative | DTNVSPNQINDTNVSPN<br>DPNANPNV--DPNANPN | Junctional |  |  | 1 |
| PF3D7_0826100 | HECT-like E3 ubiquitin ligase, putative | IQNNTNPNYE<br>VDPNANPNVD | Junctional |  |  | 1 |
| PF3D7_0827600 | conserved Plasmodium protein, unknown function | MNEFNY<br>LNELNY | N-terminus |  |  | 1 |
| PF3D7_1007700 | AP2 domain transcription factor AP2-I | HANANANP<br>NANPNANP | NANP |  |  | 1 |
| PF3D7_1122900 | dynein heavy chain, putative | DSSPNRSPSKVNQ<br>NANPNANPNKNNQ | CRR end |  |  | 1 |
| PF3D7_1346400 | VPS13 domain-containing protein, putative | NINPNI<br>NANPNV | Junctional/NVDP |  |  | 1 |
| PF3D7_1475400 | cysteine repeat modular protein 4 | NVNPNTQ<br>NANPNAN | NANP |  |  | 1 |

\*Alignment shown for each target protein (top) against CSP (bottom). Junctional, Junctional region between region 1 and the central repeat region (CRR), NVDP, minor repeat motif within the central repeat region (CRR); NANP, major repeat motif within the CRR; CRR end, end of the CRR; C-terminal, beginning of the C-terminal region (N-terminal to the Th2R epitope); N-terminus, beginning of the N-terminal region.

### Monoclonal antibodies targeting CSP can bind to a wide range of off-target *P. falciparum* proteins in a cross-reactive manner

To address the possibility that we are observing separate antibodies binding to multiple targets, protein and peptide microarrays were probed with monoclonal antibodies mAb4 and mAb10, and monoclonal antibody CIS43 was probed on protein microarrays. All three mAbs are known to block sporozoite invasion of hepatocytes. While mAb4 and mAb10 primarily bind the major repeat region (NANP), CIS43 binds both the major repeat region and the junctional region (25). The mAbs bound multiple proteins on the protein microarray at a high signal-to-noise ratio (normalized signal intensity ≥2), confirming that the monoclonal antibodies are poly-reactive. mAb4, mAb10, and CIS43 bound to 11, 142, and 6 proteins, respectively (**Fig. 6A, Fig. S8**), and each of them bound to CSP, but not necessarily to the other set of proteins hypothesized to be cross-reactive (**Fig. 6B**). mAb4 and mAb10 both bound to LRR9, but only mAb10 bound to MSP5, CLAMP, GSK3, PLP3, RZnFing and SURF8.1 (**Table S6**). CIS43 bound to CSP and only one other significantly immunogenic antigen, PF3D7_0210200, a conserved protein of unknown function. To demonstrate that mAb binding was specific and not a result of non-specific binding to the Fc region or otherwise, a well-characterized mAb against the HIV-1 gp120 protein, ARP-12033, was probed on the Pf whole proteome protein microarray, and no proteins were serorecognized (**Fig. S9A**), but gp120 and gp140 proteins were recognized on pan-HIV protein microarrays (**Fig. S9B**).

**Fig. 6.**
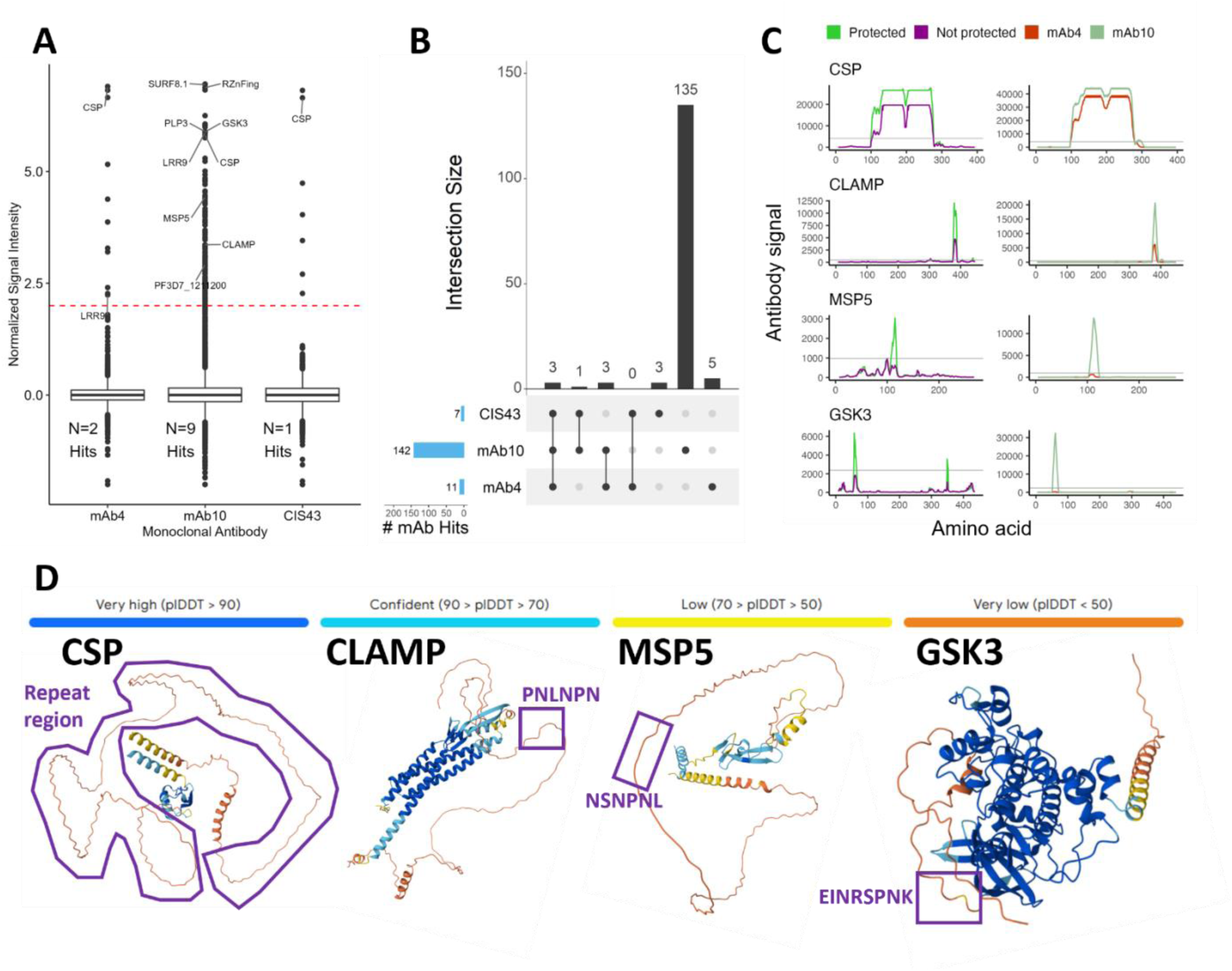
Monoclonal antibodies against the CSP bind to varying numbers of off-target *P. falciparum* proteins. **(A)** The boxplot shows the normalized signal intensity (SI) distribution for monoclonal antibodies mAb4, mAb10 and CIS43 probed on Pf whole proteome protein microarrays. A subset of proteins identified in the analysis of serum sample responses correlated with protection (Fig. 3) are labeled, as well as the number of serum sample serorecognized proteins (“hits”) for each monoclonal antibody (SI≥2.0, the red dashed line). **(B)** The upset plot shows the overlap in hits between the 3 monoclonal antibodies. Blue horizontal bars show the total number of hits for each monoclonal antibody, and vertical bars show the number of hits overlapping at specific intersections, which are shown in the dot map. **(C)** The peptide microarray profiles show the comparison between the serum medians of protected (green) and not protected (purple) individuals on the left vs. the profiles of mAb4 (orange) and mAb10 (light green) on the right. The 4 rows of plots show peptide microarray antibody profiles for CSP, CLAMP, MSP5 and GSK3, respectively. **(D)** AlphaFold 3 structural predictions for CSP, CLAMP, MSP5, and GSK3 with level of confidence depicted by blue, cyan, yellow, and orange shading and putative antibody-binding epitopes marked with purple outlines and annotated with the non-CSP NANP-like sequences observed as peptide microarray peaks.

For antibody responses on the peptide microarrays, we defined putative epitopes as those with antibody binding above mAb-specific serorecognition cutoffs for at least 4 consecutive amino acids. Amongst 131 Pf proteins probed with mAb4 and mAb10 on a peptide microarray, mAb4 bound 19 proteins at 30 putative epitopes and mAb10 bound 37 proteins at 52 putative epitopes. BLAST-p was used for pairwise alignment of the putative epitopes with the amino acid sequence of CSP and identified 6 and 10 proteins with homology to CSP for mAb4 and mAb10, respectively. Overall, the binding targets of mAb4 and mAb10 had some overlap with the putative epitopes of non-CSP proteins that were identified by the PfSPZ Vaccine samples in protein and peptide microarrays. Moreover, these shared targets have homology to CSP epitopes, re-enforcing our finding that off-target, poly-reactive antibodies could be elicited by the CSP on the surface of the PfSPZ Vaccine sporozoites. Antibody responses to CSP, CLAMP, MSP5 and GSK3 peptides are shown in **Fig. 6C**. CLAMP was serorecognized by the PfSPZ Vaccinees and had antibody peaks for both monoclonal antibodies, notably at lower intensity for mAb4 than mAb10. MSP5 was identified by the PfSPZ Vaccinees serum response and mAb10 but not mAb4, as seen with the protein microarray. CLAMP and MSP5 were both noted at the outset of the current investigation as having potential cross-reactive epitopes (15). The putative epitope of GSK3, EINRSPNK, did not have homology with CSP, but did bind mAb10, suggesting that the poly-reactivity could be against a conformational epitope that does not have linear sequence homology with CSP. A summary of putative cross-reactive CSP epitopes and the locations to which they mapped are presented in **Table 2**.

To explore conformational epitope binding, we visualized de novo amino acid sequence modeling of CSP (as a comparator), and CLAMP, MSP5, and GSK3 using AlphaFold 3 (AF3) (26) **(Fig. 6D)**. As has been predicted and observed in other structural studies of CSP (27, 28), the CSP major repeat region was predicted as a region of very low confidence (plDDT < 50), i.e., an intrinsically disordered region. The AF3 predictions of CLAMP, MSP5 and GSK3 also had regions of very low confidence, and the mAb10 binding epitopes identified by peptide microarrays fell within these unstructured regions.

### Anti-CSP monoclonal antibodies bind to proteins in an unrelated organism, *Neisseria meningitidis*

Following the observation that mAb4, mAb10 and CIS43 bind to unrelated Pf proteins, we sought to explore binding to unrelated proteins beyond *Plasmodium*. *Neisseria meningitidis* (Nm) proteome microarrays, which were available and had been tested for meningococcal mAb binding, were used to probe the Pf mAbs. Like Pf binding profiles, mAb4 and mAb10 both bound to multiple Nm proteins, while CIS43 bound to one Nm protein (**Fig. 7A, Table S7**). There was little overlap of recognized Nm proteins between the mAbs, with just one protein bound by both mAb4 and mAb10 (**Fig. 7B**). Epitope homology analysis identified 11 Nm proteins with aligned NANP/NVDP-like sequences (**Table S8**). To again ensure that mAb binding was specific, the ARP-12033 mAb against the HIV-1 gp120 protein was probed on the Nm protein microarray and showed no serorecognition or binding to Nm proteins (**Fig. S9C**).

**Fig. 7.**
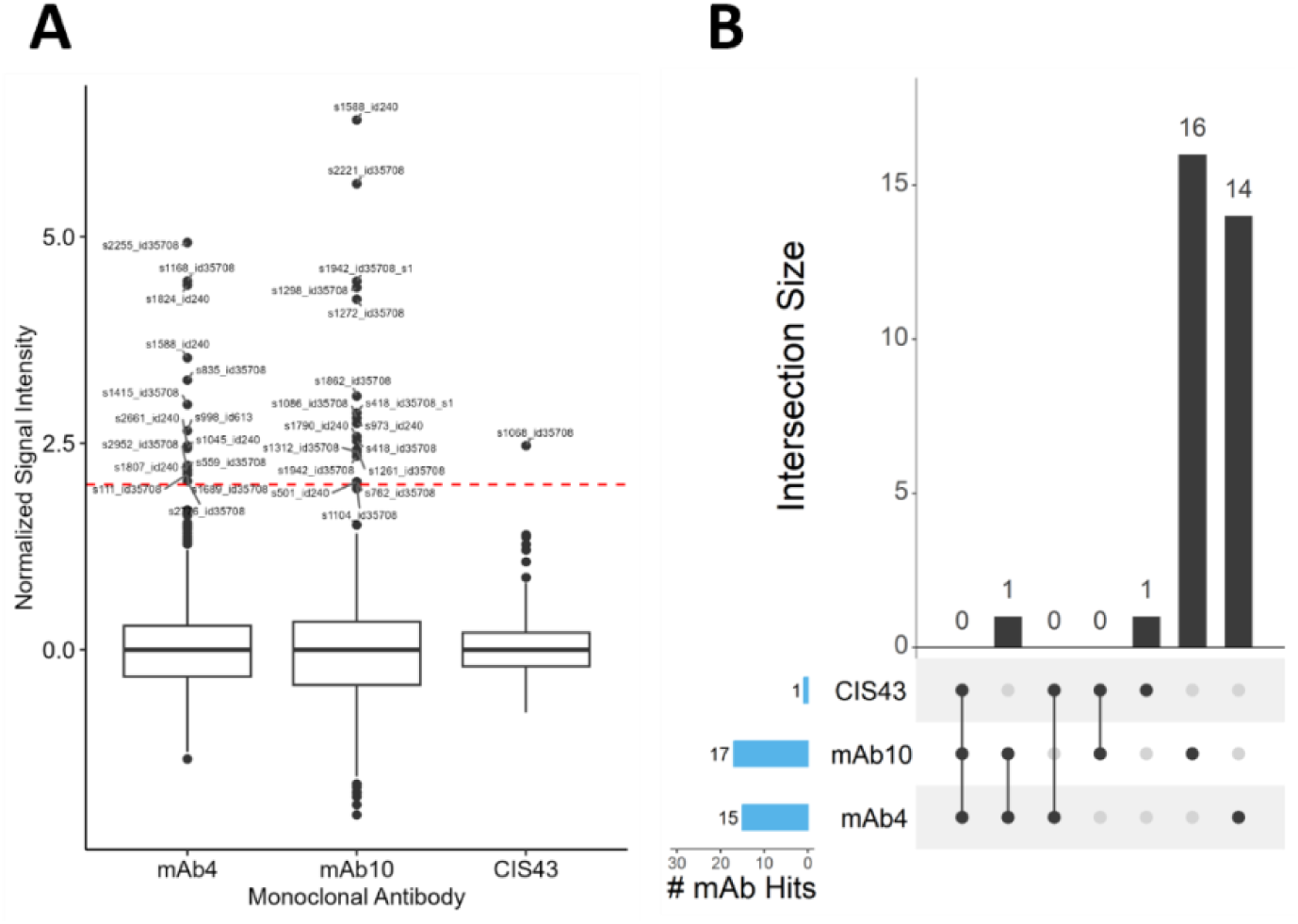
Monoclonal antibodies against CSP bind to *Neisseria meningitidis* proteins on meningococcal protein microarrays. **(A)** The boxplot shows the normalized signal intensity (SI) distribution for monoclonal antibodies mAb4, mAb10 and CIS43 probed on *Neisseria meningitidis* protein microarrays. Proteins identified as significant hits (SI≥2.0, the red dashed line) are labeled. **(B)** The upset plot shows the overlap in hits between the 3 monoclonal antibodies. Blue horizontal bars show the total number of hits for each monoclonal antibody, and vertical bars show the number of hits overlapping at a specific intersection. Intersections are shown in the dot map below the grey bars.

## DISCUSSION

One of the most intriguing outcomes of this study is the evidence that anti-CSP antibodies against the central repeat region and the junctional region cross-react with numerous other Pf proteins and even proteins from a non-*Plasmodium* organism. Our use of high-density peptide microarrays allowed us to map the antibody responses to precise epitopes on the proteins identified by protein microarrays both in this study and in RTS,S-vaccinated children (15). CSP epitopes and many of the identified off-target epitopes share sequence homology, which aligns with the premise that the antibodies are cross-reactive. Because RTS,S is a CSP subunit vaccine, post-vaccination antibody responses to non-CSP Pf proteins in malaria-naïve recipients must be cross-reactive with a vaccine component. In malaria-naïve adults given RTS,S and challenged with CHMI, off-target responses were associated with protection, and peptide-level analysis localized differentially reactive regions containing sequence similarity to CSP epitopes, including regions not represented in RTS,S (16). These findings extend the association reported in RTS,S-vaccinated children, where post-vaccination antibody levels to non-CSP proteins including MSP5, GSK3, CLAMP and DOC2 were associated with protection (15). Because serum is polyclonal, binding to both CSP and non-CSP proteins cannot be distinguished from the co-occurrence of separate, protein-specific antibodies. By contrast, a single monoclonal antibody species with off-target binding is poly-reactive by definition and cannot be explained as a mixture of antibodies with different specificities. Here, CSP-specific human monoclonal antibodies isolated from memory B cells or plasmablasts from PfSPZ Vaccine trial participants bound multiple off-target Pf proteins, demonstrating poly-reactivity via NANP-like promiscuous epitopes. Our findings extend the observation of off-target cross-reactivity to recipients of a whole-organism vaccine presenting all surface-exposed antigens on the Pf sporozoite. However, the question remains: are these cross-reactive antibodies functionally distinct from non-cross-reactive antibodies? In the current study, greater antibody breadth, i.e., more cross-reactive CSP antibodies, was associated with protection, consistent with the findings from the two RTS,S cohorts (15, 16).

Our study demonstrated different degrees of cross-reactivity for three CSP mAbs. CIS43, which primarily targets the junctional region and has previously shown very high levels of *in vitro* inhibition of primary hepatocyte infection (25) and protection when administered to Malian adults before the transmission season (29), was the least cross-reactive. Between mAb4 and mAb10, which both preferentially target the major repeats but not the junctional region, mAb10 had more cross-reactive protein targets than mAb4. In previous studies, mAb10 had greater *in vitro* inhibition of primary hepatocyte infection and inhibition of liver stage invasion in a mouse model than mAb4 (25). Importantly, the mAbs tested are a tiny representation of the repertoire of CSP antibodies in circulation after immunization with PfSPZ Vaccine, thus inference on the effect of cross-reactivity with antibody function is limited.

When examining the cross-reactive protein targets themselves, predicted functions and subcellular localization varied widely. Pf merozoite surface protein 5 (MSP5) is expressed in the sporozoite, hepatic, and blood stages of the parasite life cycle and has been considered as a vaccine candidate antigen, but never developed as one (19, 20, 22). Claudin-like apicomplexan microneme protein (CLAMP) is a predicted microneme protein that may be extruded to the parasite surface during host cell invasion (30) and would be a plausible target for functional antibodies. It is also expressed during both the sporozoite and merozoite stages (31). On the other hand, glycogen synthase kinase 3 (GSK3) is a kinase with a cytoplasmic subcellular localization, which suggests it is unlikely to be surface-associated and a target of functional antibodies. Until proven otherwise, these protein targets can still be considered possible vaccine candidate antigens. Future studies can evaluate the direct effect of antibody responses to the identified targets.

One explanation for the association between antibody cross-reactivity and protection is that cross-reactive antibodies may not act directly on the recognized targets but instead be indirectly involved or co-linear but independent of the protective response. Antibody-dependent Fc-mediated effector cell activity and complement fixation are known to contribute to protection against malaria (32–34) and have been associated with RTS,S-mediated protection (35, 36). Protection could be mediated by T-cell responses initiated through Fab engagement, with antigen binding inducing Fc conformational changes that drive T-cell activation, as described by Orlandi et al (37). Increased expression of memory-activated natural killer cells (38) and Vδ2+ T cells (18) appear to be protective against malaria and are thought to occur independently of antibody responses. Future studies could explore the association of antibody cross-reactivity with Fc-mediated and other independent effector cell responses.

Another possibility is that vaccine-elicited antibody cross-reactivity serves as an indicator for the ability to bind the intrinsically disordered major repeat region of CSP, induce structural changes, and expose functional regions of the protein (28, 39). While this may appear to contradict the smokescreen hypothesis–which posits that the major repeat region elicits high titers of short-lived, non-functional antibodies during natural infection–vaccination may overcome this mechanism by inducing a more durable antibody response and/or superior memory response. Thus, the same epitope may be immunologically distracting in natural infection but protective when targeted by vaccine-elicited antibodies.

Beyond CSP, low-complexity regions across *P. falciparum* tend to be intrinsically disordered, immunodominant, and cross-reactive, which may have an evolutionary purpose rather than exist as a result of stochastic processes (40, 41). Notably, *P. falciparum* has a higher-than-average frequency of low-complexity regions (42). Our discovery of regions of cross-reactivity could be serendipitous, in that searching for NANP-like regions between targets was a logical starting point when using BLAST-p, or it may coincide with an enrichment of cross-reactive asparagine-rich low-complexity domains contained in *Plasmodium* and non-*Plasmodium* genomes. Nonetheless, our findings suggest that cross-reactivity may serve as an indicator of important low-complexity regions, providing an impetus to identify others, and drawing attention to mechanisms of plasmodial evolution that may be implicated in immune evasion. While studying antibodies associated with exposure to malaria remains valuable for identifying potential vaccine candidate antigens, focusing on post-vaccination responses may favor the identification of meaningful “uncommon immunity.” Whether other antigens that elicit immunological smokescreens could also serve as viable vaccine candidate antigens remains to be seen.

Previous peptide-level studies of malaria antibodies have confirmed known immunogenic Pf epitopes (43–46). Raghavan et al identified common motifs among different Pf seroreactive proteins that were associated with malaria exposure in a pediatric Ugandan cohort (44). However, because exposure in that study was defined using only the most recent prior infection, and the full history and genetic sequences of infections were unknown due to natural exposure, it is unclear whether the shared inter-protein motifs reflect co-reactivity (separate responses to different infections) or true cross-reactivity. Our observation of poly-reactive mAbs is stronger support that these are cross-reactive responses. In our study, the variation observed in antibody immunogenicity counts among individuals could be a function of having a higher or lower proportion of cross-reactive CSP antibodies in the B cell and plasmablast pool. As demonstrated by Raghavan et al, while repeated exposures in malaria endemic settings also elicit antibody responses derived from memory and plasmablast pools, it is difficult to distinguish which antibodies were formed previously vs. recently. Future studies of the sequential development of the plasmablast pool over repetitive infections are ongoing and could elucidate the contributions of B-cell subsets to cross-reactivity (47).

We also observed cross-reactivity across taxa between *P. falciparum* and *N. meningitidis*. The surprising finding that poly-reactive mAbs against a Pf protein cross-react with bacterial proteins raises questions about whether NANP-like sequences are prevalent throughout microbial genomes, perhaps through an intrinsic quality of asparagine and/or proline rich domains being common in low complexity regions, or if other cross-reactive motifs are also widely distributed among microbial genomes. While not analyzed specifically, we assume Pf proteomes contain a higher proportion of intrinsic disordered regions than *Neisseria*, as Eukaryota tend to have larger fractions of disordered residues than bacteria (48);s *Plasmodium* in particular is enriched for intrinsically unstructured proteins (42). Pf has a higher fraction of low complexity regions and asparagine content than Nm (20% and 14% vs. 5% and 4%, respectively), and we were unable to find evidence that *Neisseria* is an outlier compared to other bacterial proteomes (**Table S9**)(49). Despite these differences, the mAb data suggest that there may be similarity in antigenic domains across taxa. Our study highlights our incomplete knowledge of the extent to which these epitopes occur in microbial populations.

In addition to cross-reactivity, our study highlights the proteome-wide response to a metabolically active, non-replicating sporozoite vaccine. While the response to PfSPZ Vaccine is partly dominated by CSP and cross-reactive antibodies, a subset of immunogenic proteins lacked the correlation observed with the cross-reactive set of proteins. However, it was primarily the set of cross-reactive proteins that were associated with protection. An exception may be SSP2/TRAP and AMA1, which were immunogenic in the high dose IM regimen, and although the IM group was not powered to assess correlates of protection given its low efficacy, both antigens, each unrelated to CSP cross-reactivity, are vaccine candidates associated with protection from malaria in past studies (50, 51).

Our study was somewhat limited by the small sample size. Another limitation was that we were only able to observe antibody binding to linear but not conformational epitopes on peptide microarrays. Future studies could examine conformational epitopes in the context of the proteins we identified (52). While structural prediction models that use deep learning like AlphaFold show immense power and potential, modeling proteins with intrinsic disordered regions (IDRs) remains a challenge (53). As AI-based methods are advancing rapidly, there is hope that future structural prediction methods will lead to a better understanding of these IDRs and how antibodies interact with them. Despite these limitations, the discovery that protection is associated with greater levels of intra- and extra-*Plasmodium* cross-reactivity raises new questions about parasite biology and microbial evolution, including how cross-reactivity relates to functional immunogenicity and shapes antigenic diversity.

## METHODS

### Sex as a biological variable

The original studies enrolled both males and females; however, females could not be pregnant and agreed to avoid pregnancy during their study participation. In the analysis, sex was not considered a biological variable due to sample size limitations.

### Study participants

Serum samples were from three completed, open-label clinical trials of PfSPZ Vaccine in malaria-naïve North American adults as shown in **Table 1** (17, 18, 14). For the treatment arms included in the current study, all participants received 3-4 doses of PfSPZ Vaccine (5.4 x 10^5^ - 1.35 x 10^6^ total sporozoites) and received CHMI with a homologous parasite strain three weeks after the last vaccination.

### Study Design

Two independent, but complementary proteomics platforms were used for profiling IgG antibody binding to Pf proteins: protein microarrays containing full-length or fragmented proteins, and peptide microarrays containing overlapping peptides. The proteome microarray technology at Antigen Discovery, Inc. (ADI, Irvine, CA, USA) was used to fabricate whole proteome protein microarrays. The whole proteome Pf protein microarray contained 8,871 full-length or fragmented proteins representing 5,233 unique genes from the Pf3D7 genome, or approximately 99% of the Pf3D7 protein-coding genome. A partial Nm proteome protein microarray was designed by ADI and the Bidmos Laboratory at Imperial College London and contained 998 full-length or fragmented proteins representing 982 unique genes (unpublished). Nimble Therapeutics, Inc. (now part of AbbVie, Inc.) fabricated peptide microarrays. Two peptide microarrays were used, one for the PfSPZ Vaccine samples and the second for monoclonal antibodies. Proteins were represented as 16-amino acid peptides with 1-step tiling (15-amino acid overlap). For the PfSPZ Vaccine samples, the entire *P. falciparum* proteome was represented for the NF54 strain. For the monoclonal antibodies, *P. falciparum* proteins included the NF54 variants of CSP, a subset of five proteins with antibody responses that correlated with protection based on protein microarray results (PF3D7_1030200, PF3D7_0206900, PF3D7_0312400, PF3D7_1243900, PF3D7_0906700), 123 additional proteins identified by literature search as important to pre-erythrocytic immunity, and two gametocyte stage proteins. The same protein microarray platform was used in the study of RTS,S-vaccinated children (15), and the same peptide microarray platform in the study of malaria-naïve RTS,S recipients (16), allowing direct comparison of antibody binding across studies. ELISA using purified recombinant CSP and MSP5 was used to validate protein microarray results. Serum samples that were collected at baseline (pre-vaccination) and prior to CHMI (post-vaccination) were tested on protein microarrays and peptide microarrays. Three mAbs, mAb4, mAb10 and CIS43, that were previously demonstrated to bind full length CSP and the major NANP repeat were also tested (25). The CIS43 mAb also binds the junctional region proximal to the major repeat. mAb4 and mAb10 were probed on both the protein and peptide microarrays. CIS43 was probed on the protein microarray.

### Statistical Analysis

IgG binding for both protein and peptide microarrays was first characterized as positive and negative for each protein or peptide feature and then used to classify proteins as serorecognized or not-recognized. All features and serorecognized features only were visualized using unsupervised dimension reduction methods t-stochastic neighbor embedding (tSNE) and uniform manifold approximation and projection (UMAP). PfSPZ Vaccine immunogenicity (change from pre-vaccination to post-vaccination IgG binding levels) was tested on protein microarray data using linear mixed effects regression controlling for subject-level random effects and adjusting for vaccine regimen. Univariate longitudinal analysis was also performed using paired T tests and Wilcoxon’s sign rank tests. Difference between post-vaccination and pre-vaccination antibody levels (“Deltas”) was used to analyze correlates of protection by logistic regression, unpaired univariate tests (T tests and Wilcoxon’s rank sum tests), and permutation tests. Correlation of protein microarray and ELISA data was assessed using Pearson’s correlation coefficient. To assess the effect of responsiveness to multiple Pf antigens on protection, an immunogenicity score was calculated for each individual as the number of antigens for which the Delta value was greater than 1.0 (“2-fold pre-vaccination”). In addition, unsupervised clustering of Deltas was performed using hierarchical clustering, cutting the dendrogram trees into two clusters of higher and lower responders. Correlation between CSP responses and responses to non-CSP Pf proteins was performed using Pearson’s correlation coefficient. All P values were adjusted for the false discovery rate; all statistical tests were two-tailed where applicable. For peptide microarrays, differential seroreactivity for all serorecognized proteins in the protected vs. not protected cohorts was identified using permutation tests without adjustment for multiple comparisons. BLAST-p (54) and SSearch were used for pairwise alignment of putative epitopes. AlphaFold 3 was used to predict tertiary structure and visualize binding epitopes of putative cognate or cross-reactive antigens. Detailed Materials and Methods are included in the **Supplementary Materials**.

## Supporting information

PeptideArray_all_proteins

PeptideArray_all_peptides

## Study approval

The original studies were approved by the relevant institutional review boards, and written informed consent was obtained from all participants prior to participation. Evaluation of samples in the current study was either approved by the University of Maryland, Baltimore Institutional Review Board (Protocol #HP-00058155; for evaluation of samples from the VRC-314 clinical trial) or determined to be non-human studies research (all other samples).

## Author contributions

Designing research studies: JJC, JVP, AAT, PLF, SLH, AHI, FAB, RAS, KEL, AAB, CVP

Conducting experiments: ADS, JME, JVP, AAT, JCT, RSP

Acquiring data: ADS, JME, KEL, JEE Analyzing data: JJC, AO, AAB

Interpreting results: AAB, JJC, AHI, KEL, ST-H, DJF-K, RAS, SLH

Providing reagents: FAB

Writing the manuscript: JJC, AAB, AHI

All authors reviewed the manuscript and approved the final version

## NIH public access statement

This work is the result of NIH funding, in whole or in part, and is subject to the NIH Public Access Policy. Through acceptance of this federal funding, the NIH has been given a right to make the work publicly available in PubMed Central.

## Data Availability

All data produced in the present study are available upon reasonable request to the authors.

Values for all data points in graphs are reported in “Supporting Data Values.xlsx.” Sample information, *P. falciparum* and *N. meningitidis* protein microarray data, and mode-max peptide microarray values for the serorecognized proteins are provided in “Supplemental_Data.xlsx.” Peptide microarray data are provided in full as two additional files: protein-level mode-max values for all 6,060 NF54 proteins (PeptideArray_all_proteins.csv) and peptide-level log_2_ signal intensities for all 320,909 peptides (PeptideArray_all_peptides.csv.gz). Analytic code used to generate all figures and tables is available from the corresponding author upon request.

## Acknowledgements

We thank the study participants of the PfSPZ Vaccine clinical trials, without whom this study would not be possible. We thank Angela Yee and Xiaowu Liang of ADI for support in the conduct of the protein microarray studies. We thank Lixin Gao and Tint Tint Wai of Protein Potential, LLC for performance and data of ELISAs against PfCSP and PfMSP5. We are grateful to Protein Potential, LLC and Dr. Ross Coppel for providing the recombinant PfCSP and PfMSP5, respectively.

## List of Supplementary Materials

Supplemental Materials and Methods

Supplemental Figures S1 to S13

Supplemental Tables S1 to S9

Supplemental Data: Supplemental_Data.xlsx, PeptideArray_all_peptides.csv.gz, PeptideArray_all_proteins.csv

## References

1. World Health Organization. World malaria report 2025: addressing the threat of antimalarial drug resistance.. Geneva: World Health Organization; 2025.

2. Plowe CV, Alonso P, Hoffman SL. The potential role of vaccines in the elimination of falciparum malaria and the eventual eradication of malaria. J Infect Dis. 2009;200(11):1646–1649.

3. Cockburn IA, Seder RA. Malaria prevention: from immunological concepts to effective vaccines and protective antibodies. Nat Immunol. 2018;19(11):1199–1211.

4. Clyde DF, et al. Immunization of man against sporozite-induced falciparum malaria. Am J Med Sci. 1973;266(3):169–177.

5. Rieckmann KH, et al. Use of attenuated sporozoites in the immunization of human volunteers against falciparum malaria. Bull World Health Organ. 1979;57 Suppl 1(Suppl):261–265.

6. Hoffman SL, et al. Protection of Humans against Malaria by Immunization with Radiation-Attenuated *Plasmodium falciparum* Sporozoites. J Infect Dis. 2002;185(8):1155–1164.

7. Luke TC, Hoffman SL. Rationale and plans for developing a non-replicating, metabolically active, radiation-attenuated *Plasmodium falciparum* sporozoite vaccine. J Exp Biol. 2003;206(21):3803–3808.

8. Richie TL, et al. Sporozoite immunization: innovative translational science to support the fight against malaria. Expert Rev Vaccines. 2023;22(1):964–1007.

9. Sissoko MS, et al. Safety and efficacy of a three-dose regimen of Plasmodium falciparum sporozoite vaccine in adults during an intense malaria transmission season in Mali: a randomised, controlled phase 1 trial. Lancet Infect Dis. 2022;22(3):377–389.

10. Diawara H, et al. Safety and efficacy of PfSPZ Vaccine against malaria in healthy adults and women anticipating pregnancy in Mali: two randomised, double-blind, placebo-controlled, phase 1 and 2 trials. Lancet Infect Dis. 2024;24(12):1366–1382.

11. Oneko M, et al. Safety, immunogenicity and efficacy of PfSPZ Vaccine against malaria in infants in western Kenya: a double-blind, randomized, placebo-controlled phase 2 trial. Nat Med. 2021;27(9):1636–1645.

12. Florens L, et al. A proteomic view of the Plasmodium falciparum life cycle. Nature. 2002;419(6906):520–526.

13. Mordmüller B, et al. Sterile protection against human malaria by chemoattenuated PfSPZ vaccine. Nature. 2017;542(7642):445–449.

14. Epstein JE, et al. Protection against Plasmodium falciparum malaria by PfSPZ Vaccine. JCI Insight. 2017;2(1):e89154.

15. Macià D, et al. Strong off-target antibody reactivity to malarial antigens induced by RTS,S/AS01E vaccination is associated with protection. JCI Insight. [published online ahead of print: April 21, 2022]. 10.1172/jci.insight.158030.

16. Friedman-Klabanoff DJ, et al. RTS,S-induced off-target antibody responses in malaria-naïve adults associated with protection against controlled human malaria infection. npj Vaccines. [published online ahead of print: October 24, 2026]. 10.1038/s41541-026-01600-y.

17. Seder RA, et al. Protection against malaria by intravenous immunization with a nonreplicating sporozoite vaccine. Science. 2013;341(6152):1359–1365.

18. Ishizuka AS, et al. Protection against malaria at 1 year and immune correlates following PfSPZ vaccination. Nat Med. 2016;22(6):614–623.

19. Goschnick MW, et al. Merozoite surface protein 4/5 provides protection against lethal challenge with a heterologous malaria parasite strain. Infect Immun. 2004;72(10):5840–5849.

20. Kedzierski L, et al. Immunization with a combination of merozoite surface proteins 4/5 and 1 enhances protection against lethal challenge with Plasmodium yoelii. Infect Immun. 2002;70(12):6606–6613.

21. Perraut R, et al. Association of antibody responses to the conserved Plasmodium falciparum merozoite surface protein 5 with protection against clinical malaria. PloS One. 2014;9(7):e101737.

22. Wilson KL, et al. A Synthetic Nanoparticle Based Vaccine Approach Targeting MSP4/5 Is Immunogenic and Induces Moderate Protection Against Murine Blood-Stage Malaria. Front Immunol. 2019;10:331.

23. Woodberry T, et al. Antibodies to Plasmodium falciparum and Plasmodium vivax merozoite surface protein 5 in Indonesia: species-specific and cross-reactive responses. J Infect Dis. 2008;198(1):134–142.

24. Dobaño C, et al. RTS,S/AS01E immunization increases antibody responses to vaccine-unrelated Plasmodium falciparum antigens associated with protection against clinical malaria in African children: a case-control study. BMC Med. 2019;17(1):157.

25. Kisalu NK, et al. A human monoclonal antibody prevents malaria infection by targeting a new site of vulnerability on the parasite. Nat Med. 2018;24(4):408–416.

26. Abramson J, et al. Accurate structure prediction of biomolecular interactions with AlphaFold 3. Nature. 2024;630(8016):493–500.

27. Guy AJ, et al. Insights into the Immunological Properties of Intrinsically Disordered Malaria Proteins Using Proteome Scale Predictions. PLOS ONE. 2015;10(10):e0141729.

28. Oyen D, et al. Structural basis for antibody recognition of the NANP repeats in Plasmodium falciparum circumsporozoite protein. Proc Natl Acad Sci U S A. 2017;114(48):E10438–E10445.

29. Kayentao K, et al. Safety and Efficacy of a Monoclonal Antibody against Malaria in Mali. N Engl J Med. 2022;387(20):1833–1842.

30. Sidik SM, et al. A Genome-wide CRISPR Screen in Toxoplasma Identifies Essential Apicomplexan Genes. Cell. 2016;166(6):1423–1435.e12.

31. Aurrecoechea C, et al. PlasmoDB: a functional genomic database for malaria parasites. Nucleic Acids Res. 2009;37(Database issue):D539–43.

32. Osier FH, et al. Opsonic phagocytosis of Plasmodium falciparum merozoites: mechanism in human immunity and a correlate of protection against malaria. BMC Med. 2014;12:108.

33. Lu LL, et al. Beyond binding: antibody effector functions in infectious diseases. Nat Rev Immunol. 2018;18(1):46–61.

34. Nkumama IN, et al. Breadth of Fc-mediated effector function correlates with clinical immunity following human malaria challenge. Immunity. 2024;57(6):1215–1224.e6.

35. Kurtovic L, et al. Multifunctional Antibodies Are Induced by the RTS,S Malaria Vaccine and Associated With Protection in a Phase 1/2a Trial. J Infect Dis. 2021;224(7):1128–1138.

36. Suscovich TJ, et al. Mapping functional humoral correlates of protection against malaria challenge following RTS,S/AS01 vaccination. Sci Transl Med. 2020;12(553):eabb4757.

37. Orlandi C, et al. Antigen-Induced Allosteric Changes in a Human IgG1 Fc Increase Low-Affinity Fcγ Receptor Binding. Structure. 2020;28(5):516–527.e5.

38. Loiseau C, et al. A novel population of memory-activated natural killer cells associated with low parasitaemia in *Plasmodium falciparum*-exposed sickle-cell trait children. Clin Transl Immunol. 2020;9(4):e1125.

39. Wang LT, et al. Monoclonal antibodies to the circumsporozoite proteins as an emerging tool for malaria prevention. Nat Immunol. 2024;25(9):1530–1545.

40. Ardeshir F, et al. Cross-reactive asparagine-rich determinants shared between several blood-stage antigens of Plasmodium falciparum and the circumsporozoite protein. Mol Biochem Parasitol. 1990;40(1):113–128.

41. Muralidharan V, Goldberg DE. Asparagine repeats in Plasmodium falciparum proteins: good for nothing? PLoS Pathog. 2013;9(8):e1003488.

42. Feng Z-P, et al. Abundance of intrinsically unstructured proteins in P. falciparum and other apicomplexan parasite proteomes. Mol Biochem Parasitol. 2006;150(2):256–267.

43. Jaenisch T, et al. High-density Peptide Arrays Help to Identify Linear Immunogenic B-cell Epitopes in Individuals Naturally Exposed to Malaria Infection. Mol Cell Proteomics. 2019;18(4):642–656.

44. Raghavan M, et al. Antibodies to repeat-containing antigens in Plasmodium falciparum are exposure-dependent and short-lived in children in natural malaria infections. eLife. 2023;12:e81401.

45. Friedman-Klabanoff DJ, et al. Epitope-specific antibody responses to a Plasmodium falciparum subunit vaccine target in a malaria-endemic population. J Infect Dis. 2021;223(11):1943–1947.

46. Friedman-Klabanoff DJ, et al. Controlled human malaria infection with NF54 and 7G8 strains elicit differential antibody responses to Plasmodium falciparum peptides. Front Immunol. 2025;16:1641280.

47. Ferrer P, et al. Repeat controlled human Plasmodium falciparum infections delay bloodstream patency and reduce symptoms. Nat Commun. 2024;15(1):5194.

48. Lobanov MY, Galzitskaya OV. How Common Is Disorder? Occurrence of Disordered Residues in Four Domains of Life. Int J Mol Sci. 2015;16(8):19490–19507.

49. Brüne D, Andrade-Navarro MA, Mier P. Proteome-wide comparison between the amino acid composition of domains and linkers. BMC Res Notes. 2018;11(1):117.

50. Ogwang C, et al. Prime-boost vaccination with chimpanzee adenovirus and modified vaccinia Ankara encoding TRAP provides partial protection against Plasmodium falciparum infection in Kenyan adults. Sci Transl Med. 2015;7(286):286re5.

51. Thera MA, et al. A field trial to assess a blood-stage malaria vaccine. N Engl J Med. 2011;365(11):1004–1013.

52. Martin GM, et al. Structural basis of epitope selectivity and potent protection from malaria by PfCSP antibody L9. Nat Commun. 2023;14(1):2815.

53. Zhao B, Ghadermarzi S, Kurgan L. Comparative evaluation of AlphaFold2 and disorder predictors for prediction of intrinsic disorder, disorder content and fully disordered proteins. Comput Struct Biotechnol J. 2023;21:3248–3258.

54. Schäffer AA, et al. Improving the accuracy of PSI-BLAST protein database searches with composition-based statistics and other refinements. Nucleic Acids Res. 2001;29(14):2994–3005.

