## Supplementary material for "Proteome-wide antibody responses to PfSPZ Vaccine include cross-reactive antibodies that correlate with protection from malaria": Supp_methods

### Supplemental Materials and Methods

#### Trial samples

Studies were designed to assess tolerability, safety, immunogenicity, and protective efficacy against CHMI administered by five bites of PfSPZ-infected *Anopheles stephensi* mosquitoes that met standard infectivity criteria *(53)*. PfSPZ Vaccine is comprised of NF54 strain parasites, and CHMI for the current study used either NF54 or 3D7 parasites, which are almost genetically identical *(54)*. Volunteers were monitored daily from days 7 - 29 or until treatment was complete and symptoms were resolved. Parasitemia was confirmed when *P. falciparum* parasitemia was detected in 2 PCR results, at which time volunteers were treated.

#### Monoclonal antibodies

Three mAbs, mAb4, mAb10 and CIS43, were isolated by Kisalu et al *(24)* from participants in the VRC312 PfSPZ Vaccine study *(16)* who were protected against CHMI. Monoclonal Anti-Human Immunodeficiency Virus Type 1 (HIV-1) gp120 Protein (VRC01, produced in vitro), ARP-12033 was obtained through BEI Resources, NIAID, NIH *(55)* and probed on the protein microarray.

#### Protein microarrays

*Proteome microarray construction*. Proteins were expressed using an *in vitro* transcription and translation (IVTT) system, the *Escherichia coli* cell-free Rapid Translation System (RTS) kit (5 Prime, Gaithersburg, MD, USA). A library of Pf (and Nm) partial or complete open reading frames (ORFs) cloned into a T7 expression vector pXI has been established at ADI. This library was created through an *in vivo* recombination cloning process with PCR-amplified Pf ORFs, and a complementary linearized expressed vector transformed into chemically competent *E. coli* was amplified by PCR and cloned into pXI vector using a high-throughput PCR recombination cloning method described elsewhere *(56)*. Each expressed protein included a 5' polyhistidine (HIS) epitope and 3' hemagglutinin (HA) epitope. After expressing the proteins according to manufacturer instructions, translated proteins were printed onto nitrocellulose-coated glass AVID slides (Grace Bio-Labs, Inc., Bend, OR, USA) using an Omni Grid Accent robotic microarray printer (Digilabs, Inc., Marlborough, MA, USA). Microarray chip printing and protein expression were quality checked by probing random slides with anti-HIS and anti-HA monoclonal antibodies with fluorescent labeling.

*Sample probing*. Serum samples were diluted 1:100 in a 3mg/mL *E. coli* lysate solution in protein arraying buffer (Maine Manufacturing, Sanford, ME, USA) and incubated at room temperature for 30 min. Chips were rehydrated in blocking buffer for 30 min. Blocking buffer was removed, and chips were probed with pre-incubated serum samples using sealed, fitted slide chambers to ensure no cross-contamination of sample between pads. Chips were incubated overnight at 4 °C with agitation. Chips were washed five times with TBS-0.05% Tween 20, followed by incubation with goat DyLight650-conjugated anti-human IgG (Fortis Life Sciences, Boston, MA, Bethyl Cat#A80-104D5) diluted 1:250 in blocking buffer at room temperature. Chips were washed three times with TBS-0.05% Tween 20, three times with TBS, and once with water. Chips were air dried by centrifugation at 1,000 x g for 4 min and scanned on a GenePix 4300 microarray scanner (Molecular Devices, San Jose, CA), and spot and background intensities were measured using an annotated grid file (.GAL). Data were exported in Microsoft

Excel. Monoclonal antibodies were diluted to 0.5µg/mL and probed as described above.

*Data preparation.* Raw spot and local background fluorescence intensities, spot annotations and sample phenotypes were imported and merged in the R statistical environment using R Studio *(57, 58)*, where all subsequent procedures were performed. Foreground spot intensities were adjusted by local background by subtraction, and negative values were converted to 1. Next, all foreground values were transformed using the base 2 logarithm ($Log_2$). The dataset was normalized to remove systematic effects by subtracting the median signal intensity of the IVTT controls for each sample. Since the IVTT control spots carry the chip, sample and batch-level systematic effects, but also antibody background activity to the IVTT system, this procedure normalizes the data and provides a relative measure of the specific antibody binding compared to the non-specific antibody binding to the IVTT controls (a.k.a. background). Normalized signal intensity values of 0.0 means that the intensity is no different than the background, and a value of 1.0 indicates a doubling with respect to background.

**Peptide Microarrays**

*Peptide microarray fabrication*. Peptide microarrays were synthesized with a Roche Sequencing Solutions Maskless Array Synthesizer (MAS) by light-directed solid-phase peptide synthesis using an amino-functionalized support (Greiner Bio-One) coupled with a 6-aminohexanoic acid linker and amino acid derivatives carrying a photosensitive 2-(2-nitrophenyl) propyloxycarbonyl (NPPOC) protection group (Orgentis Chemicals). Amino acids (final concentration 20 mM) were pre-mixed for 10 minutes in N,N- Dimethylformamide (DMF, Sigma Aldrich) with N,N,N',N'-Tetramethyl-O-(1H-benzotriazol-1- yl)-uronium-hexafluorophosphate (HBTU, Protein Technologies, Inc.; final concentration 20 mM) as an activator, 6-Chloro-1-hydroxybenzotriazole (6-Cl-HOBt, Protein Technologies, Inc.; final concentration 20 mM) to suppress racemization, and N,N-Diisopropylethylamine (DIPEA, Sigma Aldrich; final concentration 31mM) as base. Activated amino acids were then coupled to the array surface for 3 minutes. Following each coupling step, the microarrays were washed with N-methyl-2- pyrrolidone (NMP, VWR International), and site-specific cleavage of the NPPOC protection group was accomplished by irradiation of an image created by a Digital Micro-Mirror Device (Texas Instruments, SXGA+ resolution), projecting 365 nm wavelength light. Coupling cycles were repeated to synthesize the full in silico-generated peptide library.

*Peptide microarray sample probing*. Prior to sample binding, final removal of side-chain protecting groups was performed in 95% trifluoroacetic acid (TFA, Sigma Aldrich) and 0.5% Triisopropylsilane (TIPS, TCI Chemicals) for 30 minutes. Arrays were incubated two times in methanol for 30 seconds and rinsed four times with reagent-grade water (Ricca Chemical Co.). Arrays were washed for 1 minute in TBST (1× TBS, 0.05% Tween-20), washed twice for 1 minute in TBS, and exposed to a final wash for 30 seconds in reagent-grade water.

Serum samples were diluted 1:100 in binding buffer (0.01M Tris-Cl, pH 7.4, 1% alkali-soluble casein, 0.05% Tween-20). The monoclonal antibodies, which had an initial starting concentration of 1 mg/mL, were each diluted 1:500 and 1:1000 in binding buffer. All samples were applied to arrays and incubated overnight at 4°C. After sample binding, the arrays were washed three times in wash buffer (1× TBS, 0.05% Tween-20), 10 minutes per wash. Primary sample binding was detected via Alexa Fluor® 647-conjugated goat anti-human IgG secondary antibody (Jackson ImmunoResearch). The secondary antibody was diluted 1:10,000 (final concentration 0.1 ng/µl) in secondary binding buffer (1x TBS, 1% alkali-soluble casein, 0.05%

Tween-20). Arrays were incubated with secondary antibody for three hours at room temperature, then washed three times in wash buffer (10 minutes per wash), washed for 30 seconds in reagent-grade water, and then dried by spinning in a microcentrifuge equipped with an array holder. Fluorescent signal of the secondary antibody was detected by scanning at 635 nm at 2 µm resolution and 15% gain, using an MS200 microarray scanner (Roche NimbleGen). Scanned array images were analyzed with proprietary Roche software to extract fluorescence intensity values for each peptide. Peptide microarray signal intensities were $\log_2$-transformed.

*Peptide microarray quality control*. Distributions of NF54 peptide signal intensities were visualized across all samples for each study using boxplots and empirical cumulative distribution function (ECDF) plots (**Fig. S10-S11**). Principal components analysis, multidimensional scaling, and hierarchical clustering were visualized to inspect for outlying samples and systematic batch effects **(Fig. S12-S13)**. No systematic batch effect or outlying samples were identified; therefore, all samples were retained for subsequent analyses.

*Sliding window approach for determination of smoothed fluorescence intensities*. Array peptides were designed to span protein sequences as overlapping 16 AA long peptides (actual range 12-16 AA) with one amino acid offset. The quantity of bound immunoglobulin G (IgG) antibodies was approximated by the measured signal intensity for each peptide. Because intensities of overlapping peptides are expected to be correlated and may contain the same binding epitope, a sliding window-based average smoothing procedure with a window size of 4 AA was used to leverage information between neighboring peptides and increase the signal-to-noise ratio as previously described *(43, 59)*. A minimum of three peptides were averaged for each position along each protein sequence.

*Determination of anchoring amino acid position for each protein*. After the smoothing procedure, each protein had signal intensities corresponding to each amino acid within that protein. For the PfSPZ Vaccine samples, in many cases, the amino acid position corresponding to the maximum signal for a protein differed between individuals and between cohorts (protected vs. not-protected). Therefore, to compare signal intensities for proteins consistently between individuals, timepoints, and cohorts, a single signal intensity representing that protein was determined for each sample at the "anchoring amino acid position", using the following procedure: for each protein, the amino acid position corresponding to the maximum smoothed intensity value for each post-vaccination protected sample was determined, and the most frequently occurring amino acid position (mode amino acid position) was designated the "anchoring amino acid position." Across all samples, the signal intensity at anchoring amino acid position was compared for each protein.

*Serorecognition threshold*. For each protein, represented by the signal intensity at the anchoring amino acid position, a cutoff was defined as the mean + 2.5 standard deviations of that protein's signal intensity amongst all pre-vaccination samples (n=42, protected and not-protected groups combined). A protein was considered "serorecognized" by an individual when the signal intensity of an individual's post-vaccination sample was greater than the cutoff. For each protein, the "serorecognition prevalence" was defined as the proportion of participants for which the post-vaccination signal intensity was above the cutoff. Proteins were defined as "serorecognized" when the serorecognition prevalence for that protein was >50%.

*Definition of putative epitopes*. For the PfSPZ Vaccine samples, putative epitopes, which appear as antibody peaks, were identified as series of at least four consecutive amino acids for which the mean amino acid signal in the protected group was greater than the serorecognition cutoff. To determine putative epitopes for the monoclonal antibodies on the 131 *P. falciparum* peptide microarray, a peak definition was constructed for each monoclonal antibody. The distribution of the maximum smoothed signal intensity values for each protein was examined to identify a cutoff point using the method described by Suvorov *(60)*. The cutoff occurred at a signal intensity of 1,587 for mAb4 and 10,740 for mAb10. Antibody peaks, and therefore putative epitopes, were defined as a series of at least four consecutive amino acids for which absolute signal intensity was $\geq$750 for mAb4 or $\geq$5000 for mAb10 (i.e., the antibody peak had signal intensity of at least half of the peak intensity corresponding to the cutoff for at least four continuous amino acids).

**ELISA**

Antibody titers against recombinant CSP (supplied by Protein Potential, Rockville, MD, USA) and recombinant PfMSP5 (supplied by Coppel Laboratory, Monash University, Australia) were assessed by enzyme-linked immunosorbent assay (ELISA) as the serum dilution at which an optical density of 1.0 was determined (OD 1.0), as previously described *(61)*. Antibody ratios were calculated as the post-vaccination OD 1.0 divided by the pre-immunization OD 1.0.

**Detailed Statistical Analysis**

*Protein microarray analysis*. Quality control plots were made after each treatment of the data, which included boxplots and density plots of probe intensity by study sample and probe type. A seropositivity threshold for serum responses was established as twice the IVTT background, or a normalized signal of 1.0. Reactive antigens were defined as those that had seropositive responses in at least 50% of any of the PfSPZ Vaccine regimens at the post-vaccination time point. Non-reactive antigens were filtered before group comparisons were performed. For mAbs, a normalized signal of 2.0, or 4-fold the background, was considered as positive antigen-binding. For unsupervised dimension reduction and projection, uniform manifold approximation and projection (UMAP) from the umap package in R *(62)* was used, as well as t-stochastic neighbor embedding (tSNE) from the Rtsne package in R *(63, 64)* varying the perplexity parameter. Longitudinal pre-immunization and post-vaccination responses and covariates were analyzed with linear mixed effects regression using the lme4 package in R *(65)*. Univariate longitudinal analysis was also performed using paired T tests and Wilcoxon's sign rank tests. Difference between post-vaccination and pre-vaccination antibody levels ("Deltas") was used to analyze correlates of protection by logistic regression, unpaired univariate tests (T tests and Wilcoxon's rank sum tests), and permutation tests. Correlation of protein microarray and ELISA data was assessed using Pearson's correlation coefficient. To assess the effect of responsiveness to multiple Pf antigens on protection, an immunogenicity score was calculated for each individual as the number of antigens for which the Delta value was greater than 1.0 ("2-fold pre-vaccination"). In addition, unsupervised clustering of Deltas was performed using hierarchical clustering, cutting the dendrogram trees into two clusters of higher and lower responders. Correlation between CSP responses and responses to non-CSP Pf proteins was performed using Pearson's correlation coefficient and the corrplot package in R *(66)*. Visualization of results was performed using R base graphics, ggplot2 *(67)* and ComplexHeatmap *(68)* packages in R. All P values were adjusted for the false discovery rate using the method described by Benjamini and Hochberg, and a finding was considered significant for Benjamini-Hochberg-adjusted (BH) P-values less than 0.05 *(69)*.

*Peptide microarray analysis.* For the PfSPZ Vaccine study, for all serorecognized proteins, differential seroreactivity in the protected vs. not protected cohorts were identified using a permutation-based approach *(70)* in which the distribution of difference in means was permuted by sampling the observed population 10,000 times with all seroreactivity values randomly assigned to the "protected" or "not-protected" groups. The null hypothesis was that, in individuals after vaccination, the difference between the mean seroreactivity for the protected group and the mean seroreactivity for the not-protected group was zero. Alpha was pre-defined at 0.05; no adjustments for multiple comparisons were made.

*BLAST-p alignment of putative epitopes with CSP*. BLAST-p *(52)* with default parameters was used for pairwise alignment of the putative epitopes, identified from antibody peak signals (above), with the amino acid sequence of CSP. Alignments with E-values < 100, a high threshold necessary for alignments of short sequences, were considered significant alignments.

*AlphaFold 3*. Amino acid sequences for putative cognate or cross-reactive antigens were input to AF3 using the AlphaFold Server (https://alphafoldserver.com). Structures were colored by confidence level. Confidence was given by plDDT score as a per-atom estimate of AF3's confidence in a structure prediction (0-100 scale). Confidence values are categorized as Very High (>90), Confident (90 ≥ plDDT > 70), Low (70 ≥ plDDT > 50) and Very Low (<50).
