## Supplementary material for "Proteome-wide antibody responses to PfSPZ Vaccine include cross-reactive antibodies that correlate with protection from malaria": Supp_figs

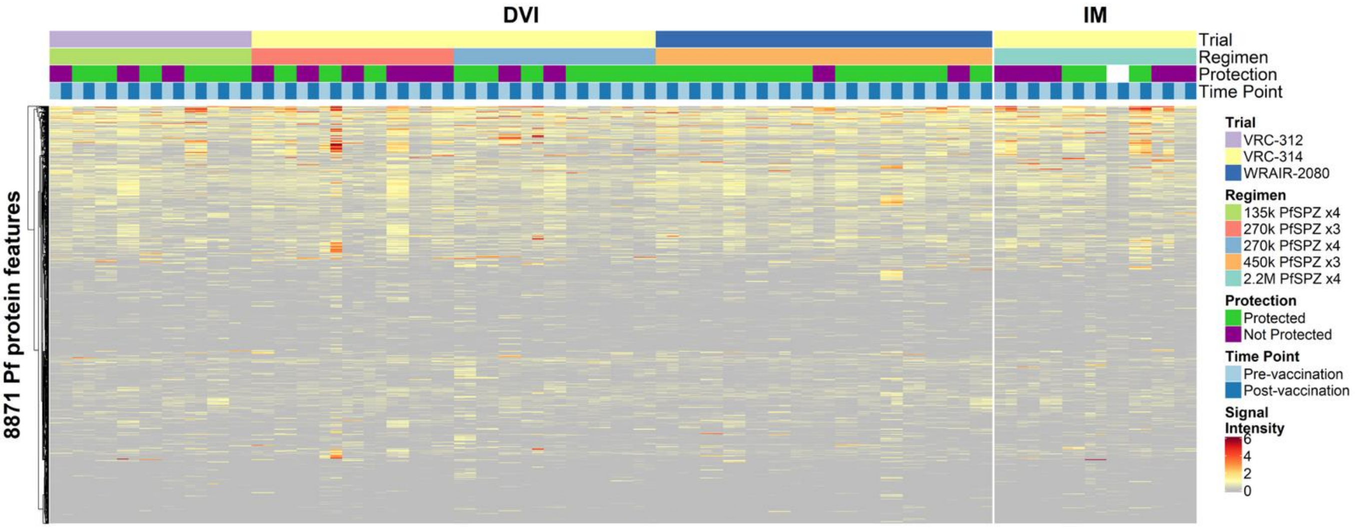


**Fig. S1. Proteome-wide antibody reactivity to PfSPZ Vaccine.** The heatmap represents normalized IgG antibody binding signal intensity for all protein features on the *P. falciparum* protein microarrays. Each row represents a protein feature, and columns represent study samples annotated with sample information above the heatmap. Samples are ordered by route of immunization, trial, and regimen. Protection or lack of protection from PfSPZ Challenge is indicated by green and purple markers, respectively, and samples from each participant are paired and shown in order from baseline to post-vaccination.

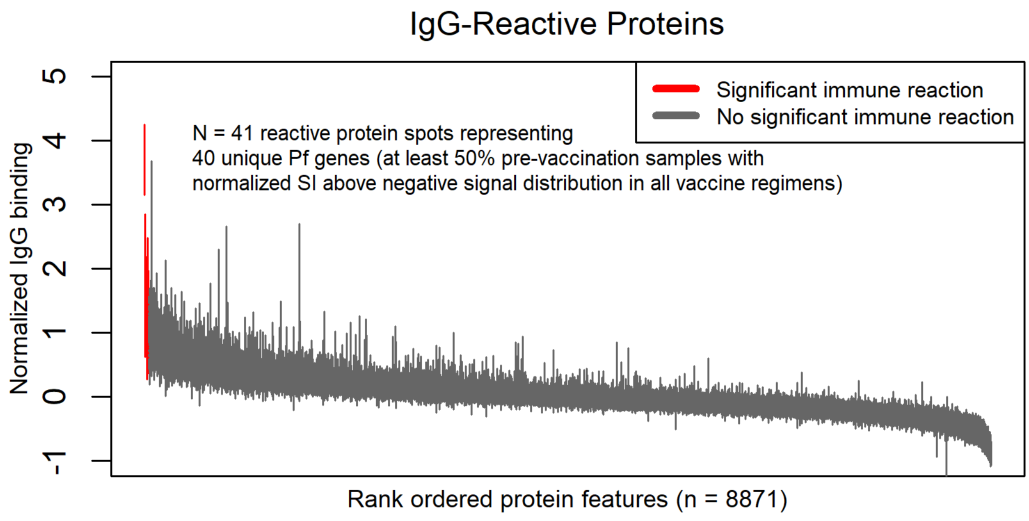


**Fig. S2. Serorecognized protein microarray protein features in pre-vaccination samples.** The error bar plot shows the interquartile range of IgG seroreactivity to each protein feature (full-length or fragmented proteins) on the *P. falciparum* protein microarrays. Protein features are ordered by median of all baseline (pre-immunization) signals, and bars are colored red if at least 50% of the study subjects had positive responses to the protein (normalized signal intensity [SI] $\geq$ 1.0).

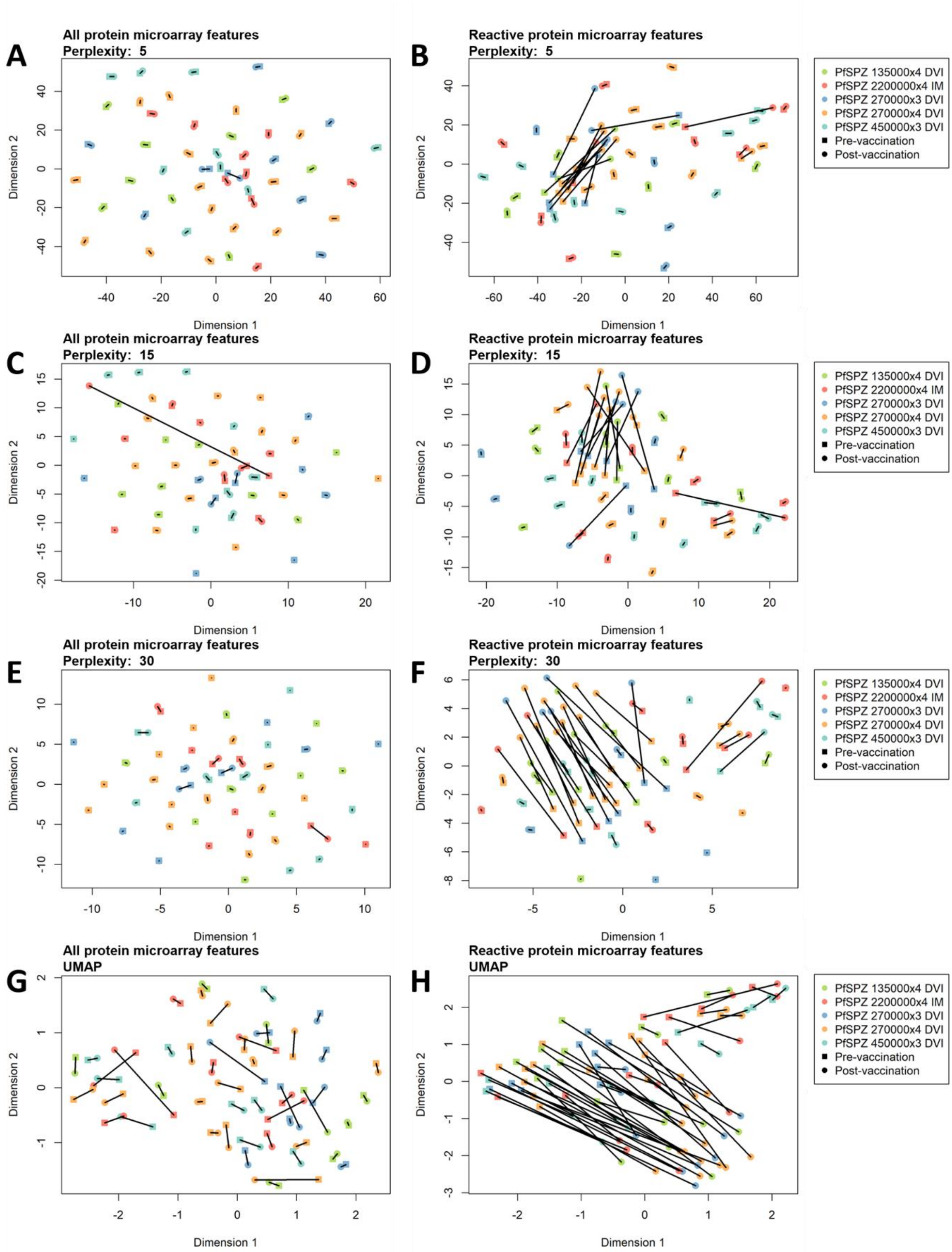


**Fig. S3. Data dimension reduction using various tSNE and UMAP models.** tSNE and UMAP were used to project the local and global structure of the protein microarray data, varying the perplexity parameter of the tSNE models. Perplexity values of 5, 15 and 30 (repeat of Fig. 1D-E parameterization) were used for both the set of all protein microarray protein features and the set of reactive antigen features. UMAP was performed using default parameters.

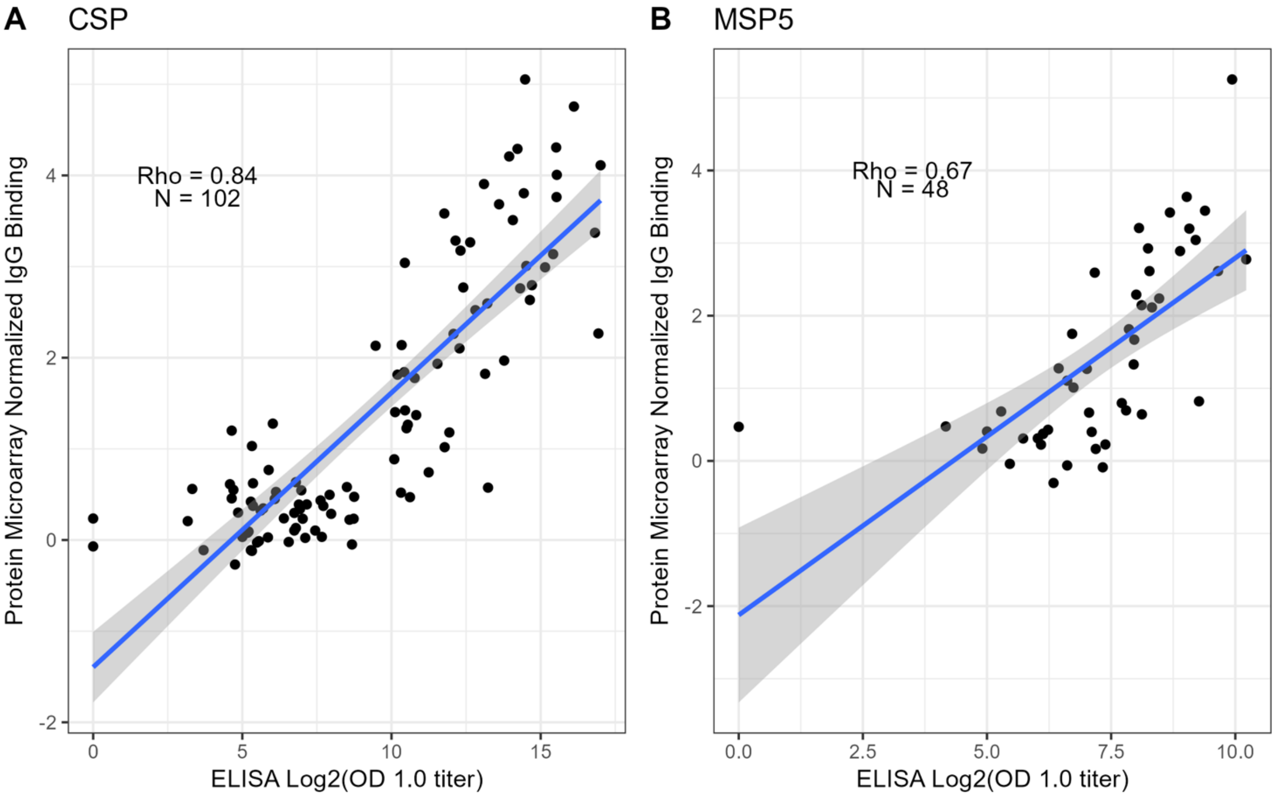


**Fig. S4. Correlation of CSP and MSP5 responses on protein microarrays vs. ELISA.** The scatter plots show the ELISA dilution factor interpolated to an optical density of 1.0 ("OD 1.0 titer") on the x axis vs. normalized signal intensity ("IgG binding") for the protein microarray of each study sample (points) for (**A**) CSP and (**B**) MSP5. Pearson's correlation coefficient ("Rho") and best fit line are shown for each antigen.

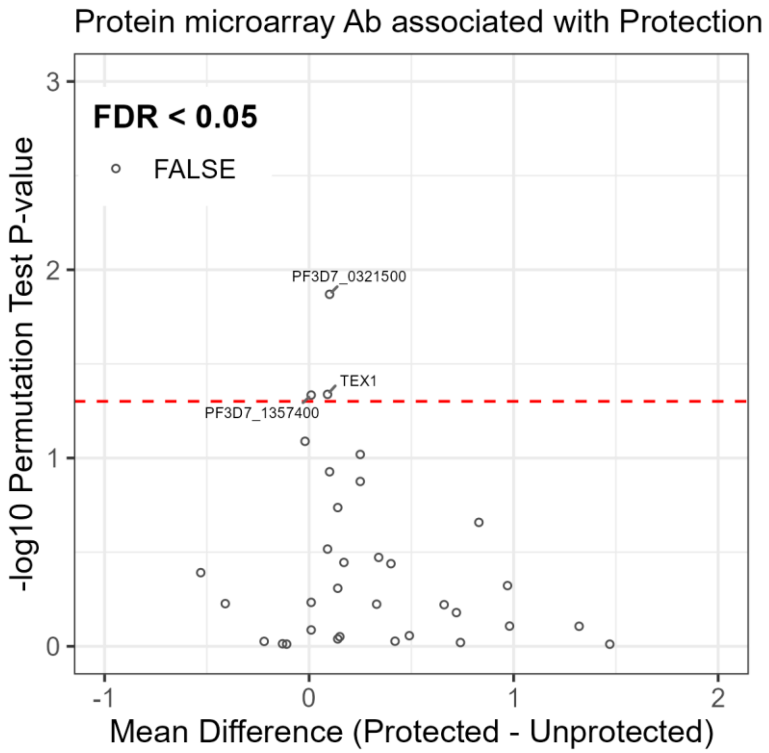


**Fig. S5. No antibody responses were associated with protection from PfSPZ Challenge in IM-immunized volunteers.** The volcano plot shows the mean difference between protected and unprotected subjects (x axis) for each of the immunogenic protein features (significantly increased IgG from baseline) from protein microarrays against the inverse $log_{10}$ P-value from permutation tests. Each point represents an immunogenic protein microarray feature; no protein features had significant responses after correction for the false discovery rate (FDR < 0.05). The red dashed line represents the unadjusted inverse log10 P-value of 0.05. Three proteins with unadjusted P<0.05 are labeled with common antigen names or gene IDs.

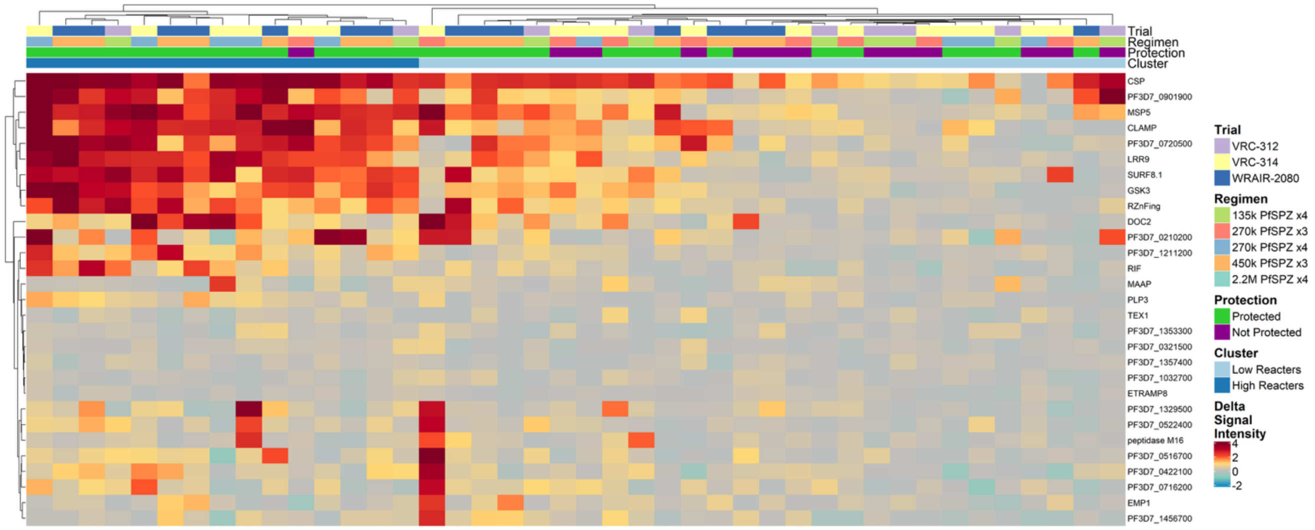


**Fig. S6. Subjects grouped as high and low reactors by hierarchical clustering and association with protection.** The heatmap represents the post- minus pre-immunization normalized antibody binding signal intensity ("Delta") for each immunogenic protein identified on the *P. falciparum* protein microarrays. For segmented proteins, the immunogenic segment with greatest statistical significance was taken to represent the protein (N=29). Each row represents a protein feature, and columns represent study samples annotated with sample information above the heatmap. Rows and columns are ordered by hierarchical clustering. The top-level branch of the column dendrogram splits the samples into two groups denoted "Cluster" that were categorized as "High" and "Low" reactors (dark blue and light blue, respectively).

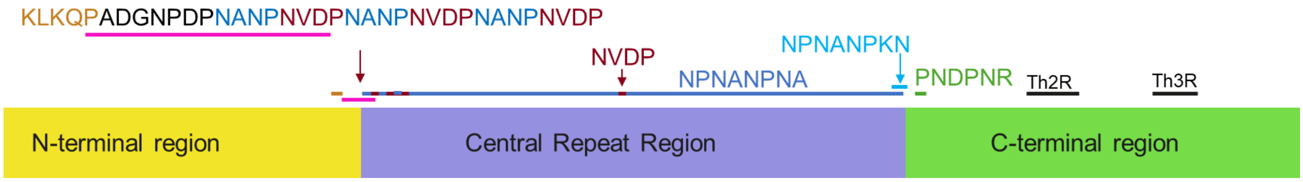


**Fig. S7. Schematic of *P. falciparum* circumsporozoite protein showing putative epitopes from literature and identified in the current study.** Schematic of *P. falciparum* circumsporozoite protein showing putative epitopes from literature and identified in the current study. Orange, junctional epitope between Region 1 (R1) and the central repeat region [8,37]. The central repeat region consists of major repeats (NPNA, blue) and minor repeats (NVDP, maroon). The putative epitopes in the current study with the strongest match (lowest E-value) aligned to the end of the central repeat region corresponding to a lysine (light blue). One putative epitope mapped to the beginning of the C-terminal region (green). Th2R and Th3R (black) are known T-cell epitopes.

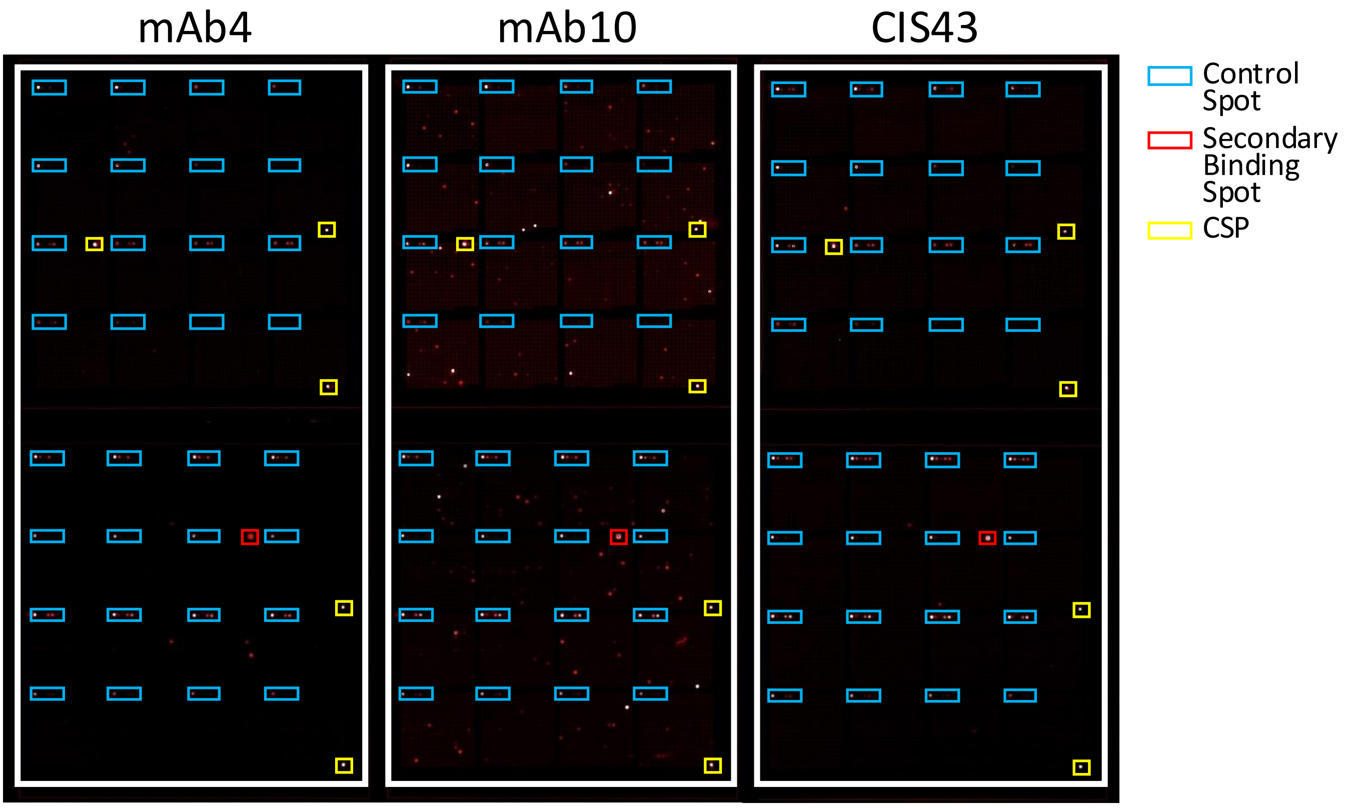


**Fig. S8. Monoclonal antibodies against the CSP bind to varying numbers of off-target *P. falciparum* proteins.** The *P. falciparum* protein microarray scanner images of mAb4, mAb10 and CIS43 show the difference in the levels of off-target binding between mAbs.

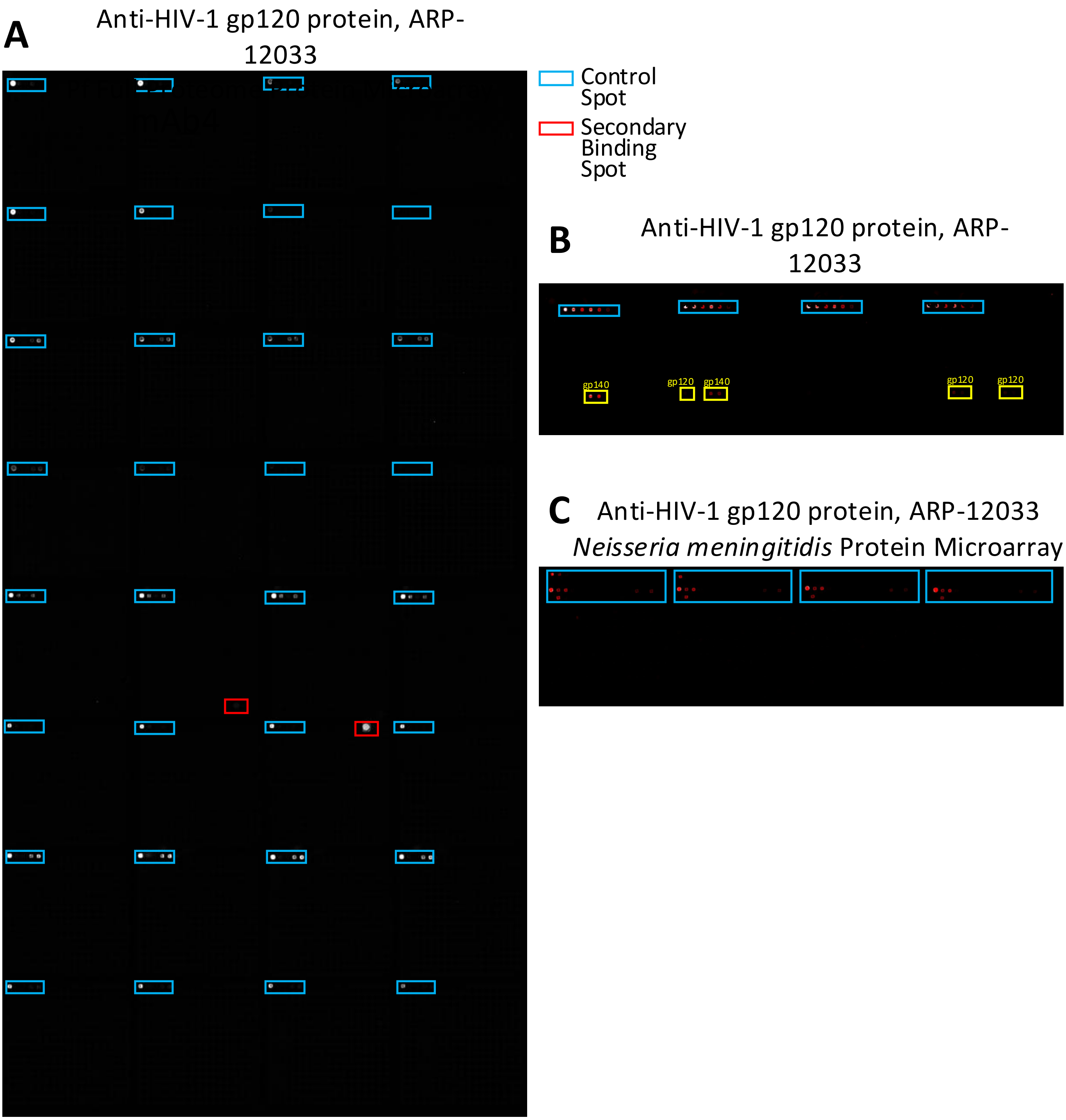


**Fig. S9. The non-malaria monoclonal antibody ARP-12033, specific for HIV gp120 protein, does not bind to off-target proteins.** The protein microarray scanner images of ARP-12033 show that it does not bind to off-target malaria proteins on the *P. falciparum* protein microarray. It binds to gp120 and gp140 proteins on the Pan-HIV protein microarray. It does not bind to off-target proteins on the *Neisseria meningitidis* protein microarray.

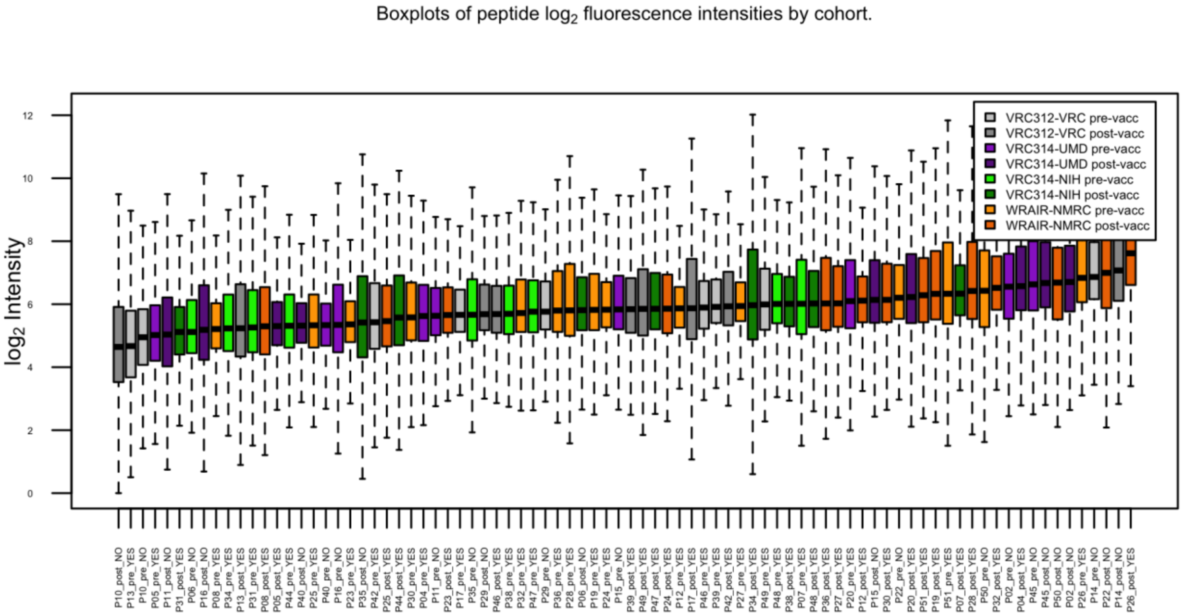


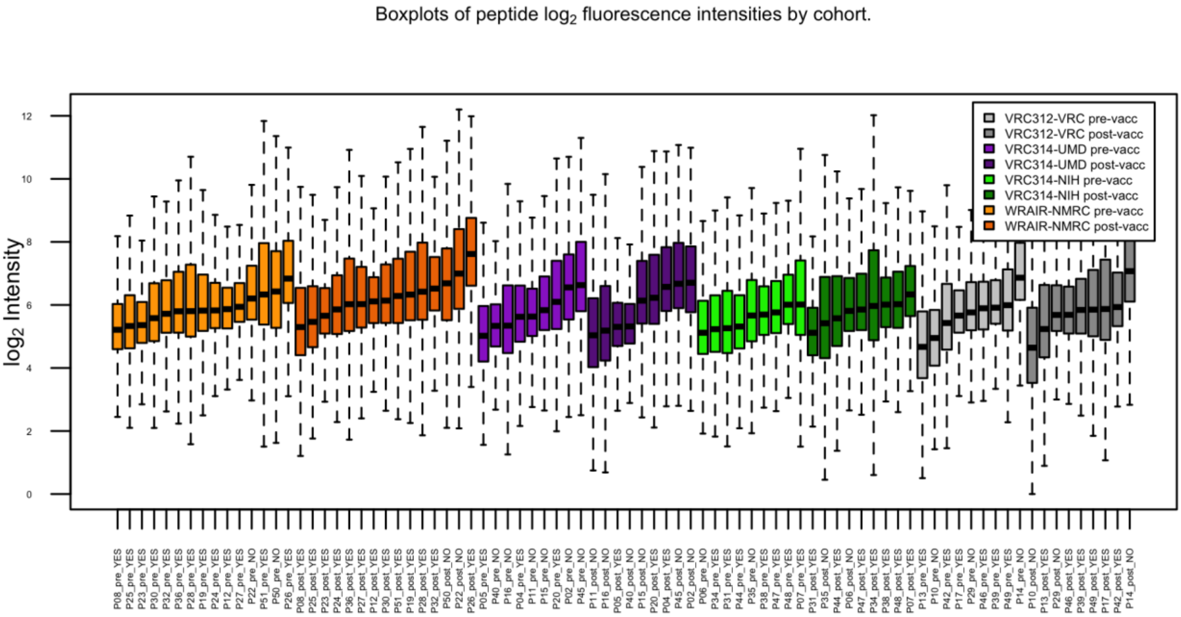


**Fig. S10. QC of peptide microarray data**. Boxplots showing distribution of peptides for each sample, arranged by ascending median value (top) and grouped by cohort (bottom).

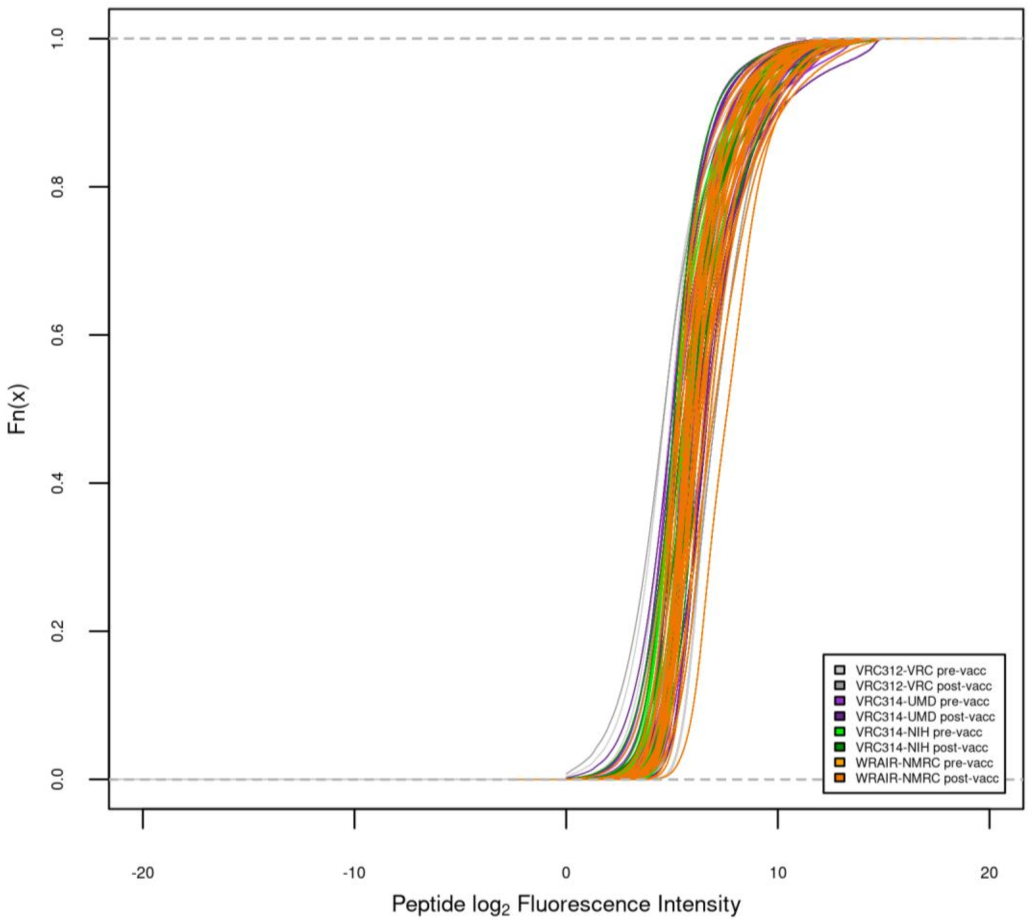


**Fig. S11. QC of peptide microarray data**. Empirical cumulative distribution function plot of peptide $\log_2$ fluorescence intensities by cohort, colored by study group and collection timepoint.

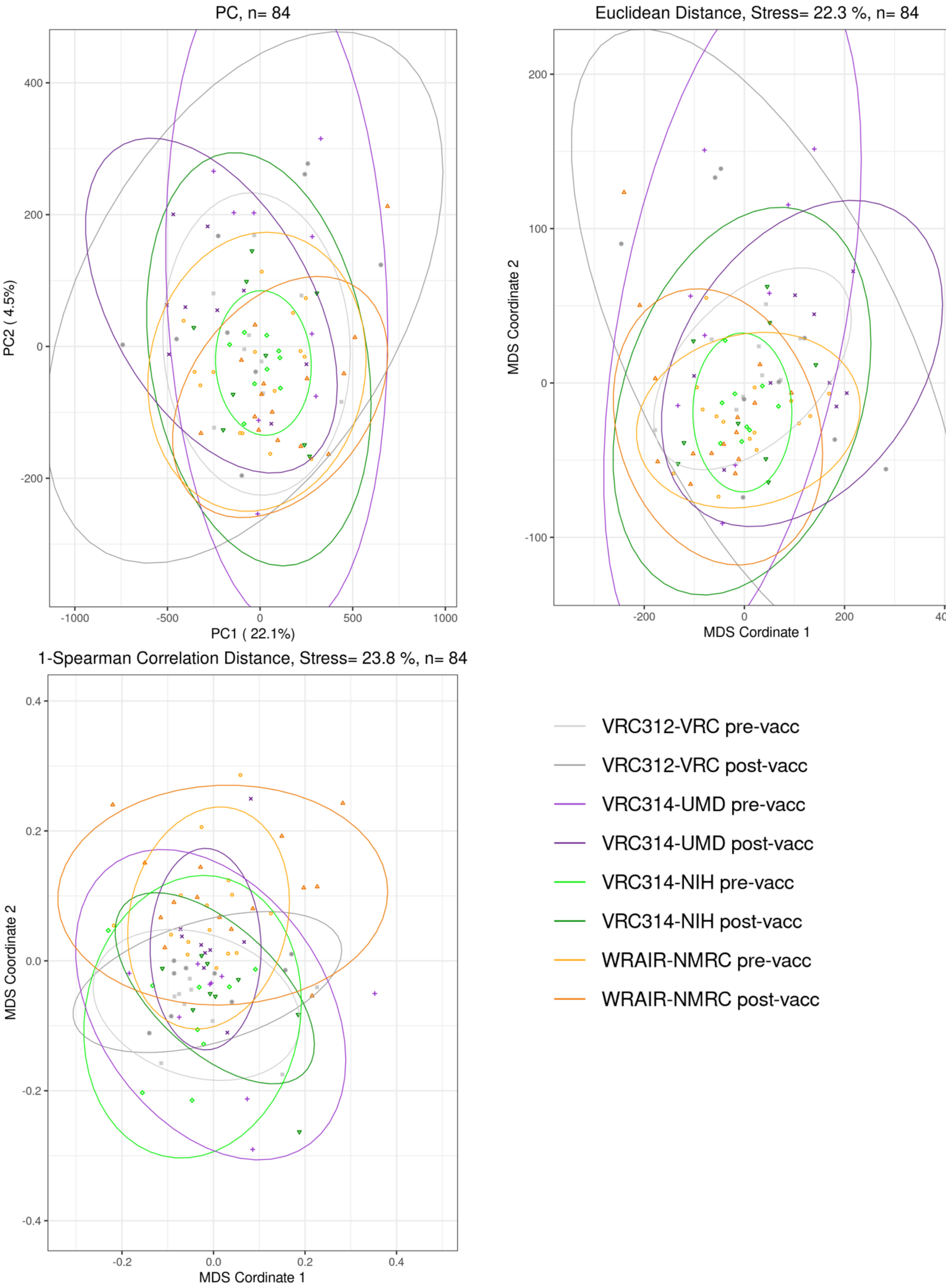


**Fig. S12. QC of peptide microarray data**. Principal components analysis and multidimensional scaling showing Euclidean distance, and 1-Spearman correlation distance, with samples colored by study group and collection timepoint.

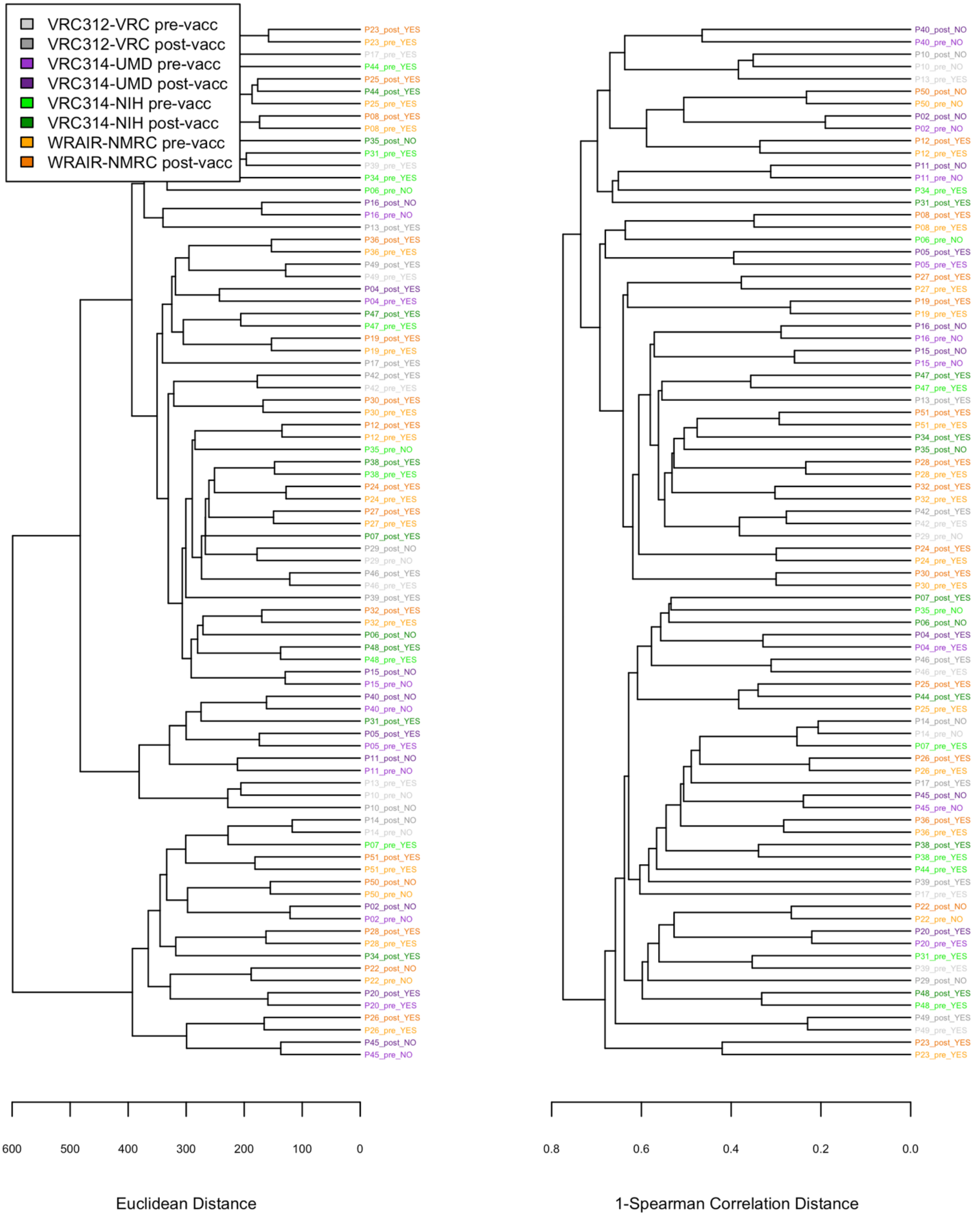


**Fig. S13. QC of peptide microarray data**. Hierarchical clustering of Euclidean distance and 1-Spearman correlation distances for samples, colored by study group and collection timepoint.
